# Seasonal Influenza Exposure Elicits Functional Antibody and T-cell Responses to A(H5) Influenza Viruses in Humans

**DOI:** 10.1101/2025.07.31.25331995

**Authors:** Mark A. Power, Willemijn F. Rijnink, Widia Soochit, Lennert Gommers, Anne van der Linden, Felicity Chandler, Femke Volker, Theo M. Bestebroer, Babs E. Verstrepen, Alba Grifoni, Ngoc H. Tan, Susanne Bogers, Gijsbert P. van Nierop, Alessandro Sette, Marion P.G. Koopmans, Corine H. GeurtsvanKessel, Reina S. Sikkema, Mathilde Richard, Rory D. de Vries

## Abstract

**Background:** Highly pathogenic avian influenza A(H5) viruses pose a pandemic threat, with a history of zoonotic spillovers into humans that are presumed immunologically naïve. Whether the general population is currently immunologically naïve to circulating A(H5) influenza viruses is unknown.

**Methods:** To evaluate the presence of cross-reactive immune responses to emerging A(H5) clade 2.3.4.4b influenza viruses in the general population, we conducted comprehensive immune profiling on cross-sectional samples from healthcare workers (n=107). Samples were collected in August and September 2024 in the scope of an ongoing prospective follow-up study: ‘Surveillance of rEspiratory viruses iN healThcare and anImal workers in the NethErLands’ (SENTINEL).

**Findings:** Low-level antibody responses directed against the A(H5) hemagglutinin (HA) head were detected in a limited number of individuals, but without hemagglutination inhibition activity. Nevertheless, we detected in most participants A(H5)-reactive antibodies with Fc-effector functions, likely directed at the conserved HA stalk. Additionally, we observed abundant neuraminidase (NA) inhibiting antibodies against avian N1s and T-cell responses against HAs and NAs from A(H5) influenza viruses. These responses correlated strongly with immune responses targeting an A(H1N1) seasonal influenza virus, indicating they were likely induced by prior exposures.

**Interpretation:** Together, our findings suggest that partial cross-reactive immunity to A(H5) influenza viruses exists in humans, which may play an important role during future outbreaks, potentially by blunting disease severity. Characterizing such baseline immunity is crucial for accurate pandemic risk assessment and preparedness planning.

**Funding:** Netherlands Organization for Health Research and Development (ZonMw), European Union’s EU4Health program DURABLE, Dutch Ministries of Agriculture, Fisheries, Food Security and Nature and Health, Welfare and Sport, National Institute of Health - National Institute of Allergies and Infectious Diseases (NIH-NIAID). The funding sources had no role in study design, data collection, analysis, interpretation of the data, or the decision to submit the paper for publication.

**Research in context:** *Evidence before this study:* We searched PubMed (Jan 1997 – Jul 2025; English) using “influenza”, “heterosubtypic”, “immunity”, “cross-immunity”, “baseline”, “population”, “H5N1”, “avian influenza”, “clade 2.3.4.4b”, “humoral”, “antibodies”, “cellular”, “T-cells”, and relevant combinations. Prior work to assess population immunity to A(H5) influenza viruses is fragmented: most reports measured binding antibodies, hemagglutination inhibition (HI) antibodies, or neutralizing antibodies exclusively, and few examined T-cell responses. The measurement of integrated antibody breadth, functional HI, neuraminidase inhibition (NI), and antibody-dependent cellular cytotoxicity (ADCC) activity, as well as T-cell responses, has not been described in cross-sectional population studies.

*Added value of this study:* We present profiling of immunity to A(H5) influenza viruses in an inferred unexposed cross-sectional sample of healthcare workers (n=107). This comprehensive analysis revealed patterns that were not detected in earlier single-endpoint studies: (i) A(H5) binding antibodies mainly targeted the conserved stalk, with higher titers in adults over 60 years, (ii) A(H5)-reactive NI-and ADCC-mediating antibodies were common, and (iii) cross-reactive T-cell responses against HAs and NAs of A(H5) influenza viruses were detected. The rational selection of antigens from both seasonal and avian influenza viruses allowed for direct comparison of multiple effector mechanisms. These design choices led to the first comprehensive map of humoral and T-cell effector functions against A(H5) influenza viruses, establishing a robust baseline for future vaccine evaluations and pandemic-risk assessments.

*Implications of all the available evidence:* Our findings in combination with existing research shows that that immunity to A(H5) influenza viruses is widespread. It is likely that repeated exposure to seasonal influenza viruses led to the development of a cross-reactive antibody and T-cell repertoire capable of recognizing emerging A(H5) influenza viruses. While the protective value of the cross-reactive immune responses remains speculative, evidence from model systems suggests these could confer protection, potentially blunting disease severity in the case of a future outbreak.

## Introduction

Highly pathogenic avian influenza A(H5) viruses represent a continuous pandemic threat. Since their emergence over two decades ago, A(H5) influenza viruses of the A/goose/Guangdong/1/1996 lineage have become established in poultry and progressively expanded in both geographic range and host species. Currently, A(H5) influenza viruses belonging to genetic clades 2.3.2.1 and 2.3.4.4 are dominating in various parts of the world. Of particular concern are the A(H5N1) clade 2.3.4.4b viruses, which have spread extensively via infected migratory birds across Europe, Asia, Africa, the Americas, and even Antarctica.^1,2^ This widespread dissemination has been accompanied by frequent spillovers into mammals,^3–9^ with the spillover into dairy cattle in the United States in 2024 being a recent example of great magnitude.^10^ For now, human A(H5) influenza virus infections remain rare, with over a thousand confirmed cases worldwide since 1997.^11–14^ However, the likely mammal-to-mammal transmission in multiple settings,^4,5,15–18^ combined with the emergence of mammalian-adaptation substitutions,^19^ heightens concerns that A(H5) clade 2.3.4.4b influenza viruses may acquire the capacity for sustained human-to-human transmission.

The rapid spread of influenza A viruses in past pandemics was primarily driven by the immunologically naïve status of the human population. During the 2009 A(H1N1) pandemic, for example, illness was predominantly observed in younger individuals with little evidence for cross-reactive antibodies. However, a subset of older adults had pre-existing immunity due to prior exposure to earlier A(H1N1) viruses, likely contributing to milder disease in that age group.^20^ Whether the general population is currently immunologically naïve to circulating A(H5) influenza viruses is unknown. However, all individuals have been exposed to seasonal influenza viruses through infection, vaccination, or a combination of both. Emerging evidence suggests that such exposure may elicit cross-reactive immune responses to A(H5) clade 2.3.4.4b influenza viruses.^21,22^ Still, the extent and functional relevance of this immunity remain incompletely understood, highlighting a critical knowledge gap.

Neutralizing antibodies against A(H5) influenza viruses, considered the most important correlate of protection against influenza, ^23^ are rarely detected in individuals who have not been exposed to this subtype. However, antibodies capable of binding hemagglutinin (HA) from various A(H5) clades, including clade 1, clade 2.1.3.2, and clade 2.3.4, are frequently observed in the general population.^24,25^ Although these antibodies do not inhibit viral entry, they can mediate effector functions through Fc-dependent mechanisms, like antibody-dependent cellular cytotoxicity (ADCC).^26–28^ In addition to antibodies, memory T-cells are thought to be important in limiting disease severity following influenza virus infection.^29–31^ Cross-reactive memory T-cells targeting conserved epitopes within the matrix (M), nucleoprotein (NP), HA, and neuraminidase (NA) proteins from A(H5) influenza viruses that circulated at the beginning of the century have been identified in small groups of healthy individuals.^32–36^ However, comprehensive cross-sectional data on binding antibodies, functional antibodies, and T-cell responses specific to circulating A(H5) clade 2.3.4.4b influenza viruses remain limited.

Collectively, prior studies have highlighted that research on cross-reactive immune responses to A(H5) influenza viruses in the general population has been fragmented, and that little to no functional data are available for A(H5) clade 2.3.4.4b influenza viruses. To support more accurate risk assessments, such as those guided by the World Health Organization (WHO) tool for influenza pandemic risk assessment (TIPRA),^37^ and inform pandemic preparedness, we investigated the presence of virus-specific binding antibodies, functional antibodies, and T-cell responses targeting A(H5) influenza viruses in a cohort of healthcare workers (HCW) in the Netherlands. This population is routinely exposed to seasonal influenza viruses through vaccination or natural infection but is unlikely to have been exposed to avian influenza viruses.

## Methods

### Study design and participants

Samples were collected at the Erasmus MC in the Netherlands in the scope of the ‘Surveillance of rEspiratory viruses iN healThcare and anImal workers in the NethErLands’ (SENTINEL) study. A detailed description of this study is reported in the **Supplemental Methods**. The study is conducted in accordance with the Declaration of Helsinki. The study protocol (NL86800.078.24) was approved by the Medical Ethics Committee of Erasmus University Medical Center; written informed consent was obtained from all study participants before the first study visit. Here, we report data obtained from a random selection of participants (n=107), from whom samples were collected during the periodic sampling in August and September 2024. Cohort characteristics are shown in **Table 1** and **Supplemental Figure 1A**.

**Table 1.**
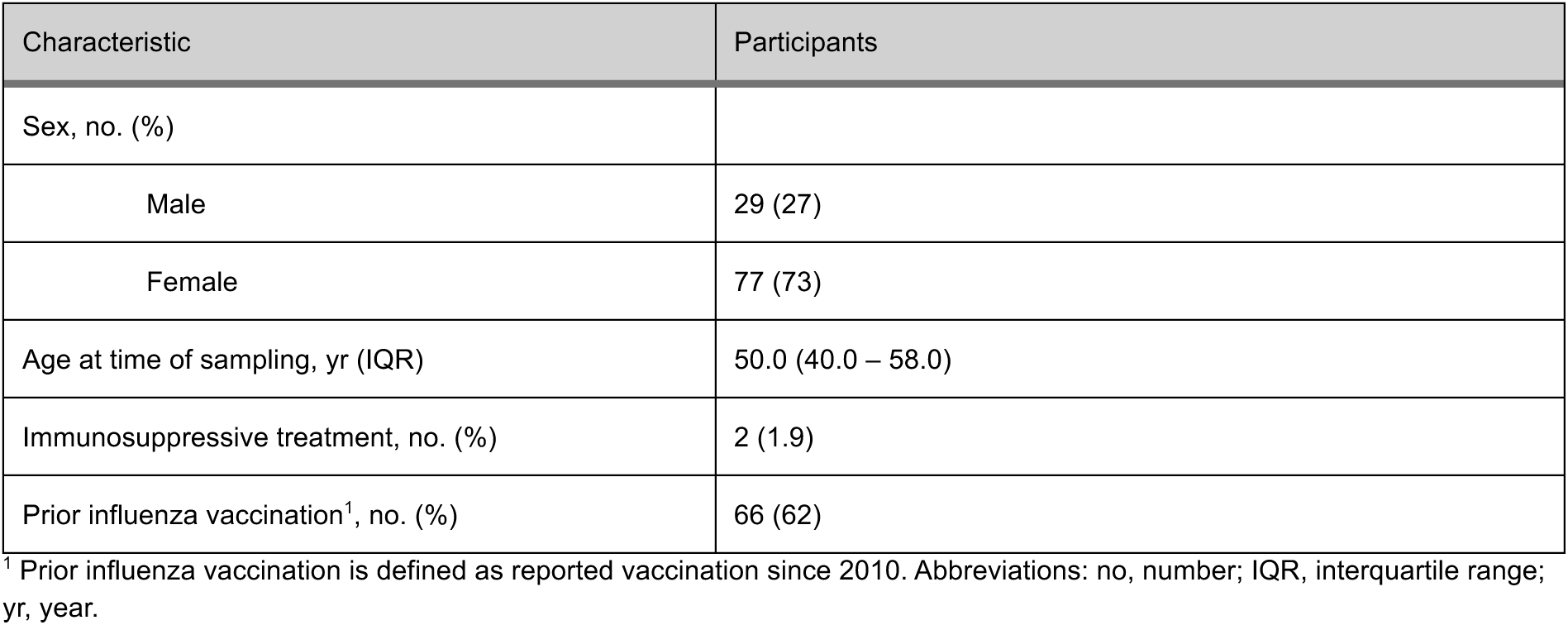
Cohort characteristics.

### Serum and peripheral blood mononuclear cells (PBMC) isolation

For serum collection, blood was collected in 10 mL tubes without anticoagulant (BD), centrifuged at 1,000 x g for 15 minutes, and serum was stored at −20°C for further experiments. Additionally, anticoagulated whole blood was collected in lithium heparin tubes (BD); a portion was used directly for T-cell assays, and PBMC were isolated from the remainder by density gradient centrifugation (details in **Supplemental Methods**). Upon use, PBMC were thawed in Iscove’s Modified Dulbecco’s Medium (IMDM; Lonza) supplemented with 10% foetal bovine serum (FCS), 100 IU/mL penicillin, 100 μg/mL streptomycin, and 2 mM L-glutamine. PBMC were treated with 50 U/mL Benzonase (Merck) for 30 minutes at 37°C prior to use in stimulation assays.

### Detection of virus-specific antibodies

Virus-specific antibodies were detected in n=106 serum samples (1 serum sample was unavailable, respective donor was only included for T-cell assays, as described below). A protein microarray (PMA) assay was performed to measure the presence of binding IgG antibodies directed to a large panel of commercially acquired HA recombinant proteins from the A(H1), A(H2), A(H3), A(H5), A(H7), and B influenza virus subtypes. HI assays were performed using wild type or recombinant viruses to evaluate the presence of functional HA-specific antibodies. The enzyme-linked lectin assay (ELLA) and neuraminidase inhibition (NI-ELLA) assay were performed with recombinant viruses to detect functional NA-specific antibodies. Fc-mediated effector functions were evaluated by the antibody-dependent cellular cytotoxicity (ADCC) assay using HA recombinant proteins and NK92.05-CD16 cells. A detailed description of the serological methods is given in the **Supplemental Methods**; a schematic overview is shown in **Supplemental Figure 1B**.

### Detection of virus-specific T-cells

Virus-specific T-cell were detected in n=107 samples. To determine T-cell response profiles, IFN-γ production was measured after stimulation of whole blood with custom-made peptide pools covering the HA or NA proteins of influenza A or B viruses of interest, the N protein of measles virus, the S protein of SARS-CoV-2 or CEFX (interferon gamma release assay [IGRA]). The activation-induced marker (AIM) assay was conducted to phenotype HA and NA-specific T-cells in a selection of IGRA high, medium, and non-responders. Cytokine detection in AIM supernatants was performed to identify cytokines released in response to HA and NA peptide stimulation using Legendplex. A detailed description of the detection of virus-specific T-cells is given in the **Supplemental Methods**; a schematic overview is shown in **Supplemental Figure 1B**.

### Statistical analysis

A detailed description of the statistical analysis plan is given in the **Supplemental methods.**

## Results

### Cohort characteristics

A total of 107 participants were included in this analysis, which was performed as part of the SENTINEL study. The cohort consisted of 77 (73%) female participants, which is in concordance with the Erasmus MC workforce; the median age was 50. Cohort characteristics are shown in **Table 1** and **Supplementary** Figure 1. Within the cohort, 66 (62%) of individuals received at least one seasonal influenza vaccination in the past 15 years, with the majority vaccinated in the last four years (**Supplemental Table 1**). Two participants were on immunosuppressive treatment (prednisolone and adalimumab, respectively).

**A(H5) influenza virus-reactive antibodies bind HA0, but not HA1.** The presence of binding IgG antibodies directed against a broad panel of HA recombinant proteins from the A(H1), A(H2), A(H3), B, A(H5), and A(H7) subtypes was determined in sera from 106 HCW by PMA (**Figure 1A, Supplemental Table 2**). HA1 subunits, mainly comprising the HA head, where the receptor binding site is located, and HA0 ectodomains, comprising the stalk and the head, were included. Explanation of virus strain abbreviations can be found in **Supplemental Table 3**. Most donors had detectable binding antibodies as determined by PMA (responder defined as binding IgG antibody titer ≥20) targeting the HA1 subunit of recent seasonal vaccine viruses: A(H1) VI/22 (49% of participants), A(H3) TH/22 (91%), B(Vic) AU/21 (96%), and B(Yam) PH/13 (100%). An age-stratified analysis revealed that older individuals had higher A(H3)-reactive IgG levels to older antigens and vice versa (**Supplemental Figure 2A**). In contrast, antibodies binding to the HA1 subunit of A(H5) influenza viruses were less frequently detected: 26% of participants for clade 2.3.4 AN/05, 7% for clade 2.3.4.4b tu/GE/16, and 12% for clade 2.3.4.4b AS/20 (**Figure 1A**). If participants had detectable antibodies, the antibody levels were low (geometric mean titer (GMT) of 13 (95% confidence interval (CI): 12-15), 11 (95% CI: 10-12), and 12 (95% CI: 11-13) for AN/05, tu/GE/16, and AS/20, respectively). When evaluating the HA0 antigens, the majority of HCW had detectable antibodies reactive with the A(H5) clade 2.3.4 AN/05 and clade 2.3.4.4b AS/20 (92% and 100%, respectively). In an age stratified analysis, antibody levels against A(H5) influenza virus HA0 antigens appeared higher in older HCW (**Supplemental Figure 2B**). This was confirmed when antibody levels were plotted per age bracket, and A(H5) influenza virus HA0 binding antibody levels appeared significantly higher in adults over 60 years of age (**Supplemental Figure 2C**).

**Figure 1.**
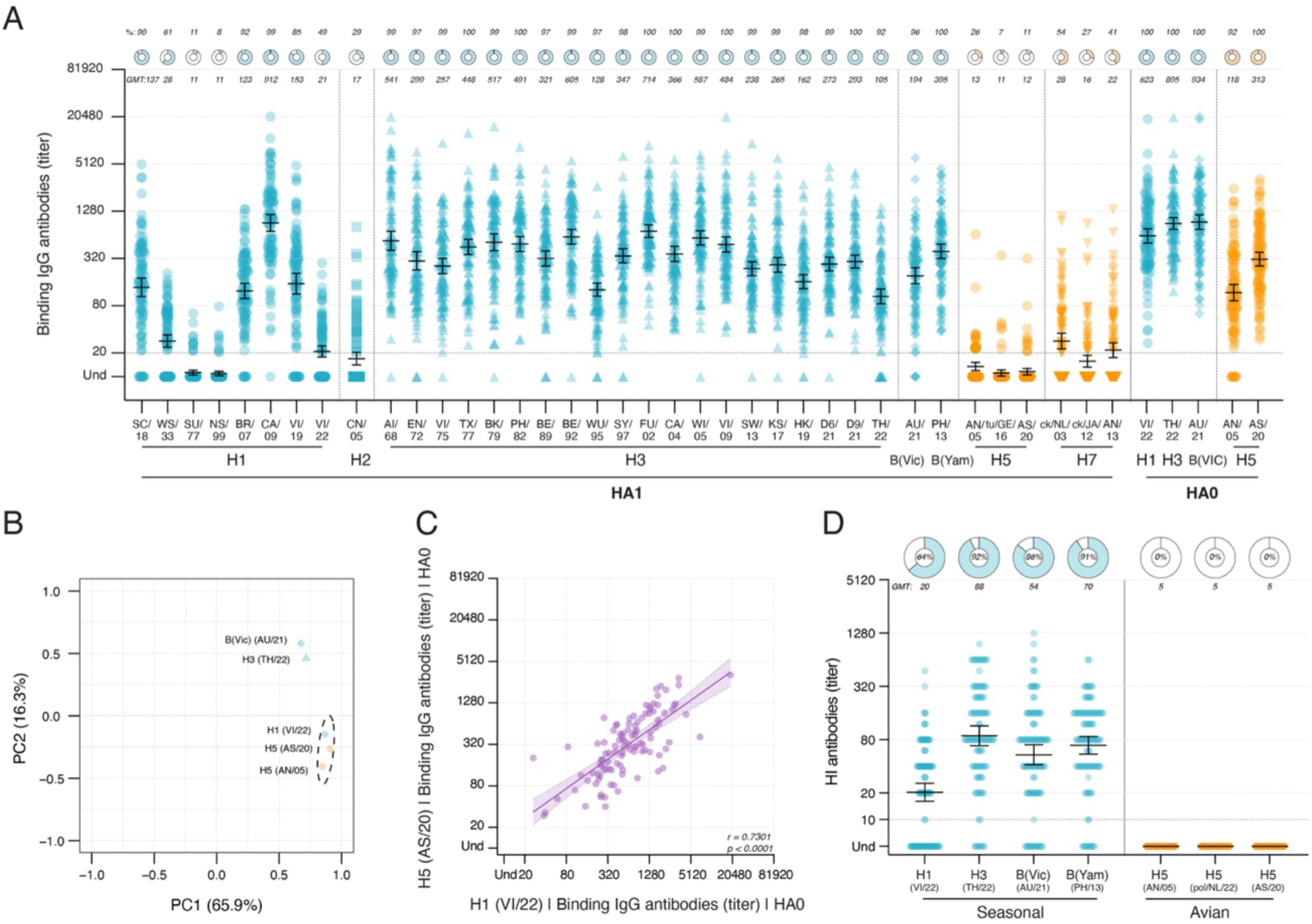
Detection of binding and HI antibodies to seasonal and avian influenza viruses. **(A)** Binding IgG antibody levels in HCW sera against a panel of HA1 and HA0 antigens from seasonal (blue, A(H1), A(H2), A(H3), B(Vic), and B(Yam)) and avian (yellow, A(H5), A(H7)) influenza viruses. The panel comprises a selection of antigens from seasonal A(H1) and A(H3) influenza viruses, dating back to 1918 and 1968, respectively, as well as recent vaccine antigens, including one A(H2) representative and B(Vic) and B(Yam) vaccine antigens. The dashed horizontal line represents the threshold titer of 20. Samples with an unmeasurable PMA titer are indicated as undetectable. **(B)** Principal component analysis (PCA) of log₁₀(x+1) transformed, then centred, and z-scaled for each antigen-specific response. PC1 and PC2 explain 65.9% and 16.3% of the variance, respectively. K-means clustering was applied to the PCA results, and a 90% confidence ellipse was computed for the primary cluster using its covariance matrix. **(C)** Scatter plot of A(H1) VI/22 versus A(H5) AS/20 HA0 binding IgG titers. The diagonal line represents the linear regression performed on log4-transformed titers. Shaded areas indicate the 95% confidence interval. Spearman correlation coefficients (r) and p-values are provided. **(D)** HI antibody responses against seasonal A(H1) VI/22, A(H3) TH/22, B(Vic) AU/21, and B(Yam) PH/13, in addition to avian A(H5) (AN/05, pol/NL/22, and AS/20) influenza viruses. The dashed horizontal line represents the threshold titer of 10. Samples with an unmeasurable HI titer are indicated as undetectable. For panels **(A)** and **(D)**, the bars indicate the GMT with a 95% confidence interval. Donut charts above each antigen show the percentage of responders (defined as binding IgG titer ≥ 20 or HI titer ≥ 10), with the percentage indicated above or within each donut chart. GMT values are listed under each donut chart. Each dot represents an individual serum sample from a total of 106 HCW. Abbreviations: HI, hemagglutination inhibition; HCW, healthcare workers; GMT, geometric mean titer; Und, undetected; PC, principal component. Virus strain abbreviations can be found in **Supplemental Table 3**.

Next, we analysed the correlation between binding IgG antibody levels to all antigens in a comprehensive matrix (**Supplemental Figure 3**). The relationship between binding IgG antibody levels to the HA0 antigens from both seasonal and avian A(H5) influenza viruses was further assessed in a dimensionality reduction principal component analysis (PCA), showing that A(H1) and A(H5) HA0 antigens clustered closely together (**Figure 1B**). In a direct correlation analysis, binding IgG antibody levels to A(H1) and A(H5) HA0 were strongly correlated (Spearman r=0.7301, **Figure 1C**).

### HI antibodies targeting A(H5) influenza viruses were absent in HCW

Since A(H5) HA0 binding antibodies were detected in HCW, but those binding the variable head domain only at low levels and in a fraction of participants, HI functionality was not expected. To confirm this, HI assays were conducted (**Supplemental Table 4**). HI antibodies targeting recent seasonal influenza viruses were highly prevalent in HCW with GMTs of 20 (95% CI: 16-26), 88 (95% CI: 68-114), 54 (95% CI: 42-70), and 70 (95% CI: 55-87), with responder rates (defined as HI titer ≥10) of 64%, 92%, 86%, and 91% for A(H1) VI/22, A(H3) TH/22, B(Vic) AU/21, and B(Yam) PH/13, respectively (**Figure 1D**). Conversely, HI antibodies targeting A(H5) influenza viruses were absent in all HCW.

### Functional ADCC responses targeting the HA0 of A(H5) influenza viruses were detected in HCW

Next, we evaluated whether the antibodies binding to the HA0 from A(H5) influenza viruses were functional through ADCC (**Supplemental Table 2**). Using NK92.05-CD16 cells, we measured natural killer (NK) cell degranulation based on CD107a surface expression following serum incubation by flow cytometry. Degranulating cells were defined as LIVE/CD56^+^/CD107a^+^ cells (**Figure 2A**). Notably, all HCW sera showed measurable ADCC activity against HA0 proteins from a seasonal and avian influenza virus, with GMTs of 298 (95% CI: 253-350) for A(H1) VI/22 and 307 (95% CI: 258-365) for A(H5) AS/20 (**Figure 2B**). The observed response to the A(H1) and A(H5) antigens was similar, as indicated by a Spearman correlation analysis (r=0.6919) (**Figure 2C**). The ADCC levels moderately correlated with binding IgG antibody levels targeting the corresponding HA0 protein as determined by PMA (Spearman r=0.4921 for A(H1) and 0.6079 for A(H5)) (**Figure 2D**). Collectively, these findings demonstrate that exposure to seasonal influenza A(H1) viruses likely induced cross-reactive antibodies targeting the HA0 of A(H5) clade 2.3.4.4b AS/20, capable of mediating functional Fc-dependent responses.

**Figure 2.**
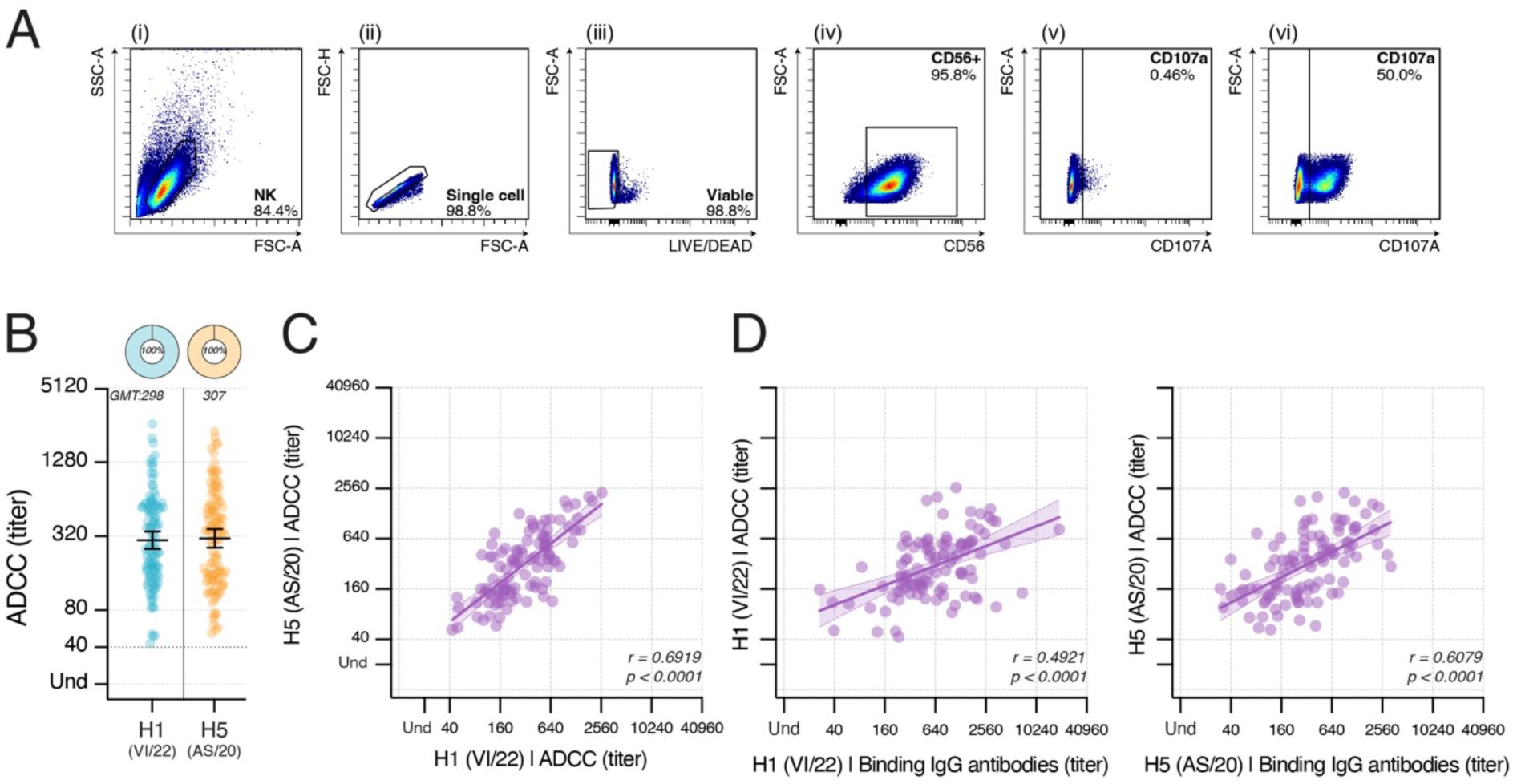
Detection of ADCC-mediating antibodies targeting seasonal A(H1) and avian A(H5) in HCW. **(A)** Gating strategy for NK92.05-CD16 degranulation: (i) NK92.05-CD16 cells are selected based on size and granularity, (ii) exclusion of doublets, (iii) exclusion of dead cells, and (iv) CD56 expression. Degranulation is measured as a percentage of CD107a^+^ NK cells (vi). Background measured on PBS-coated (v) wells is subtracted on a per-sample basis before titers are calculated. **(B)** ADCC-mediating antibody titers in HCW sera against A(H1) VI/22 (blue) and A(H5) AS/20 (yellow) HA0 antigens. The bars indicate the GMT with a 95% confidence interval. The dashed horizontal line represents the threshold titer of 40. Donut charts above each antigen show the percentage of responders (defined as ADCC titer ≥ 40), with the percentage indicated within each donut chart. GMT values for each HA0 antigen are listed under the donut chart. **(C)** Scatter plot of A(H1) VI/22 versus A(H5) AS/20 HA HA0 ADCC titers. **(D)** Scatter plots of A(H1) VI/22 HA HA0 ADCC titer versus binding IgG antibody titers (left), and A(H5) AS/20 HA HA0 ADCC titer versus binding IgG antibody titers (right). The diagonal line in each scatter plot represents the linear regression on log4-transformed titers. Shaded areas indicate the 95% confidence interval. Spearman correlation coefficients (r) and p-values are provided. For all panels, each dot represents an individual serum sample from a total of 106 serum samples. Abbreviations: HCW, healthcare workers; PBS, phosphate-buffered saline; GMT, geometric mean titer; ADCC, antibody-dependent cellular cytotoxicity. Virus strain abbreviations can be found in **Supplemental Table 3**.

### Neuraminidase-inhibiting antibodies cross-reactive with N1s from A(H5) influenza viruses were detected in HCW

Next, the presence of functional antibodies targeting N1 VI/22 and N2 TH/22 of seasonal influenza viruses, and N1 (AN/05 and pol/NL/22), N6 HU/21, and N8 AS/20 of A(H5) influenza viruses was assessed in a NI-ELLA assay (**Supplemental Table 4**). Robust inhibition of seasonal N1 and N2, with GMTs of 282 (95% CI: 210-380) and 40 (95% CI: 32-49), respectively, in addition to high responder levels (defined as a NI titer ≥10; 99% and 94%, respectively) were detected (**Figure 3A**). Notably, high antibody levels and responder rates were also observed for the two avian N1s AN/05 and pol/NL/22 with GMTs of 190 (95% CI: 147-246) and 208 (95% CI: 1531-284), in addition to responder rates of 97% and 94%, respectively. In contrast, responses to avian N6 and N8 were markedly lower with GMTs of 6 (95% CI: 5.6-7.1) and 13 (95% CI: 10-17), and responder rates of 20% and 50%, respectively. NI titers to N1s from seasonal and A(H5) influenza viruses were strongly correlated, with a Spearman r=0.6307 and 0.7296 between seasonal N1 and avian N1 AN/05 or pol/NL/22, respectively (**Figure 3B**). This correlation suggests that the presence of avian N1-specific antibodies is probably the result of past exposure to seasonal influenza A(H1N1) viruses.

**Figure 3.**
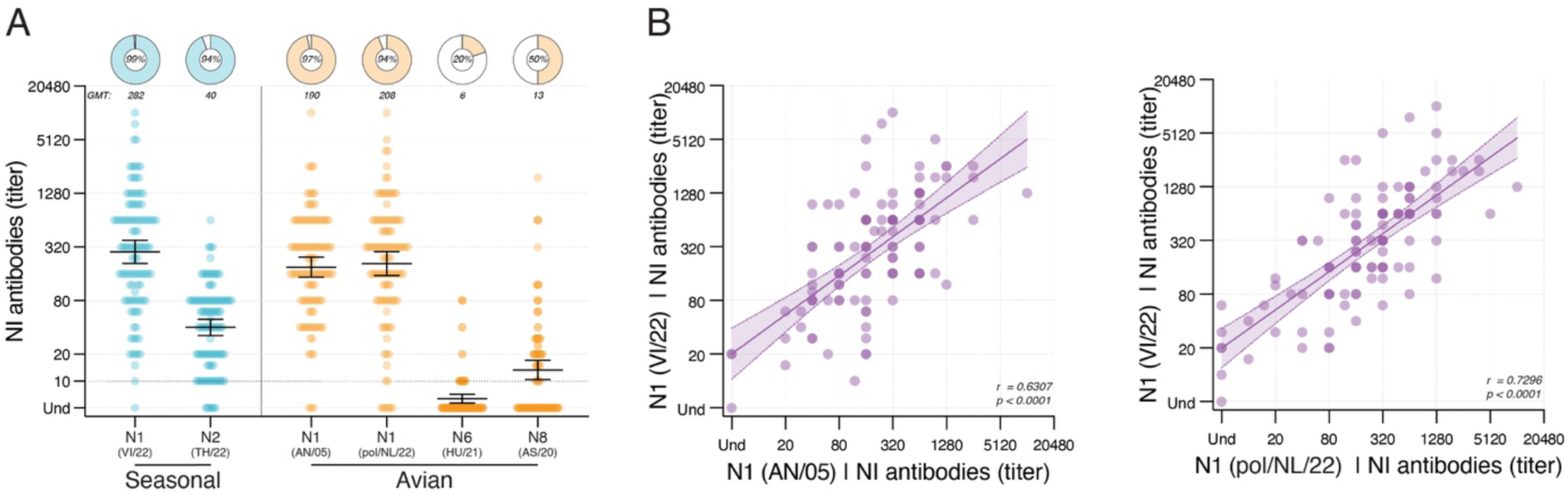
Detection of NI antibodies to NAs from seasonal and A(H5) influenza viruses. **(A)** NI antibody responses against N1 VI/22 and N2 TH/22 of seasonal influenza viruses (blue), and N1s AN/05 and pol/NL/22, N6 HU/21, and N8 AS/20 of A(H5) influenza viruses (yellow) are shown. The bars indicate the GMT with a 95% confidence interval. The dashed horizontal line represents the threshold titer of 10. Samples with unmeasurable NI titers are indicated as undetectable. Donut charts above each NA-subtype show the percentage of responders (defined as NI titer ≥ 10), with the percentage indicated within each donut chart. GMT values for each NA antigen are listed under the donut chart. **(B)** Scatter plots of N1 VI/22 versus N1 AN/05 (left) or N1 pol/NL/22 (right). The diagonal line in each correlation plot represents the linear regression on log2-transformed titers. Shaded areas indicate the 95% confidence interval. Spearman’s correlation coefficients (r) and p-values are shown. For all panels, each dot represents an individual serum sample measurement from a total of 106 serum samples. Abbreviations: NI, neuraminidase inhibition; GMT, geometric mean titer; HCW, healthcare workers; Und, undetected. Virus strain abbreviations can be found in **Supplemental Table 3**.

### Virus-specific T-cells targeting seasonal and avian HA and NA were detected in HCW

IGRAs using whole blood stimulated with overlapping peptide pools that span the HA and NA proteins from several seasonal and A(H5) influenza viruses were performed. The selected antigens were identical to those selected for the HI and NI assays (**Supplemental Table 5** and **6**). Significant recall of T-cell responses (defined as *p*<0.05 when comparing a specific antigen with the DMSO control by the Wilcoxon signed-rank test, **Supplemental Figure 4**) was observed for all antigens from influenza viruses, both human and avian. After correction for the DMSO background, T-cell responder rates (defined as >0.01 IU/mL) for human antigens were 74% for A(H1) VI/22 (geometric mean (GM): 0.06 IU/mL), 79% for A(H3) TH/22 (GM: 0.07 IU/mL), 95% for B(Vic) AU/21 (GM: 0.27 IU/mL), 97% for B(Yam) PH/13 (GM: 0.24 IU/mL), 84% for N1 VI/22 (GM: 0.07 IU/mL), and 74% for N2 TH/22 (GM: 0.04 IU/mL) (**Figure 4**). For avian antigens, T-cell responder rates were 48% for A(H5) AN/05 (GM: 0.02 IU/mL), 64% for A(H5) AS/20 (GM: 0.03 IU/mL), 69% for N1 AN/05 (GM: 0.05 IU/mL), 63% for N1 pol/NL/22 (GM: 0.05 IU/mL), 43% for N6 HU/21 (GM: 0.02 IU/mL), and 53% for N8 AS/20 (GM: 0.03 IU/mL). Data from the control conditions were as expected, with low T-cell responses targeting measles virus (indicating past infection or vaccination), and high T-cell responses to SARS-CoV-2 S (indicating recent exposure) as well as the immunodominant CEFX positive control.

**Figure 4.**
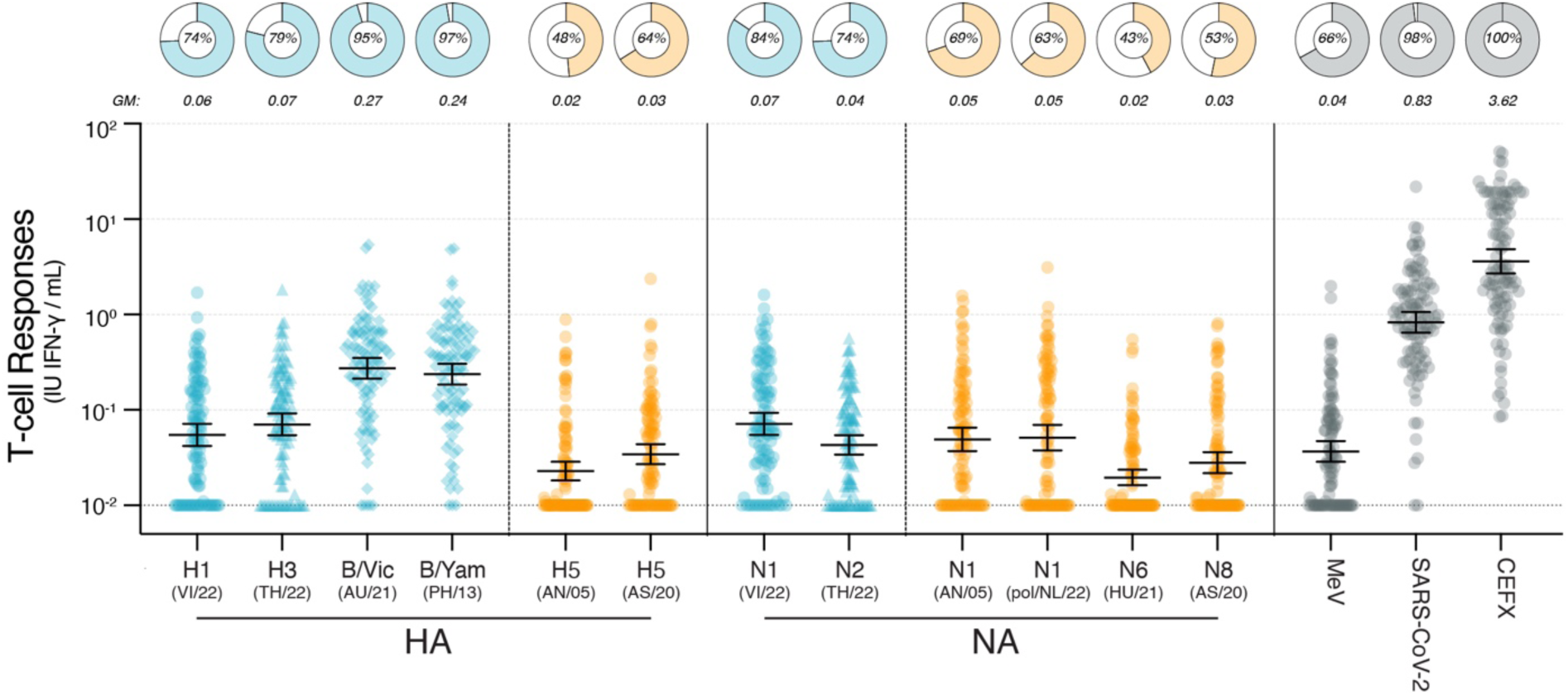
Detection of antigen-specific T-cells against HA and NA antigens from seasonal and A(H5) influenza viruses by IGRA and AIM. IFN-γ levels (IU/mL) following stimulation of whole blood with overlapping HA or NA peptide pools representative of seasonal and A(H5) influenza viruses. Measles virus N, SARS-CoV-2 S, and CEFX were included as controls. IFN-γ levels were corrected by subtraction of background values obtained upon DMSO stimulation. Uncorrected levels are shown in **Supplemental** Figure 4. The bars indicate the GM with a 95% confidence interval; the black dashed line indicates the limit of detection at 0.01 IU/mL. Donut charts above each peptide pool show the percentage of responders (defined as IFN-γ concentration > 0.01 IU/mL), with the percentage indicated within each donut chart. GM values for each peptide pool are listed under the donut chart. Each dot represents an individual sample measurement from a total of 107 samples. Abbreviations: IGRA, interferon-gamma release assay; AIM, activation-induced marker; IFN-γ, interferon gamma; IU, international units; MeV, measles virus; SARS-CoV-2, severe acute respiratory syndrome coronavirus-2; DMSO, dimethyl sulfoxide; GM, geometric mean. Virus strain abbreviations can be found in **Supplemental Table 3**.

### T-cell reactivity was confirmed by AIM flow cytometry

To further characterize antigen-specific T-cell responses, we selected PBMCs from 15 HCW characterized as high, medium, and non-responders in the IGRA assay. We assessed T-cell reactivity after stimulating PBMCs with peptide pools and detected antigen-specific cells using AIM. Antigen-specific activated CD4^+^ and CD8^+^ memory T-cells were identified by exclusion of naïve cells (CD45RA^+^/CCR7^+^), and subsequent selection of those CD4^+^ and CD8^+^ T-cells that co-expressed the activation markers CD137^+^/CD134^+^ and CD137^+^/CD69^+^, respectively (**Figure 5A**). After subtracting the DMSO background, antigen-specific CD4^+^ and CD8^+^ memory T-cells targeting seasonal A(H1) VI/22 and N1 VI/22, and avian A(H5) AS/20 and N1 pol/NL/22 influenza antigens were detected in the majority of HCW (**Figure 5B and 5C**), irrespective of the IGRA stratification. Next, cytokine secretion profiles in AIM supernatants were assessed (**Supplemental Figure 5**). These secretion profiles mostly confirmed the AIM data; however, in supernatants from some samples that tested negative in AIM, virus-specific production of cytokines could still be detected, apart from one donor, indicative of low-frequency T-cell responses in all samples. Cytokines related to antigen-specific Th1 cells (IL-2, IFN-γ, and TNF-α) were most abundantly detected, with IL-2 found in all but one supernatant. Antigen-specific Th2/Th9 (IL-4, IL-5, IL-9, IL-10, and IL-13) cells were also observed in most supernatants. AIM^+^ CD4 frequencies and IL-2, IFN-γ, and IL-5 secretion in response to A(H5) peptide pool stimulation were lower than in response to A(H1) peptide pool stimulation, indicative of a cross-reactive response (**Figure 5B** and **Supplemental Figure 5)**. IFN-γ levels in AIM supernatants correlated significantly with IFN-γ levels measured in IGRA, demonstrating the robustness of both assays (Spearman r=0.6643, **Supplemental Figure 6**). Collectively, these results highlight the presence of cross-reactive T-cells targeting A(H5) influenza virus antigens in HCW.

**Figure 5.**
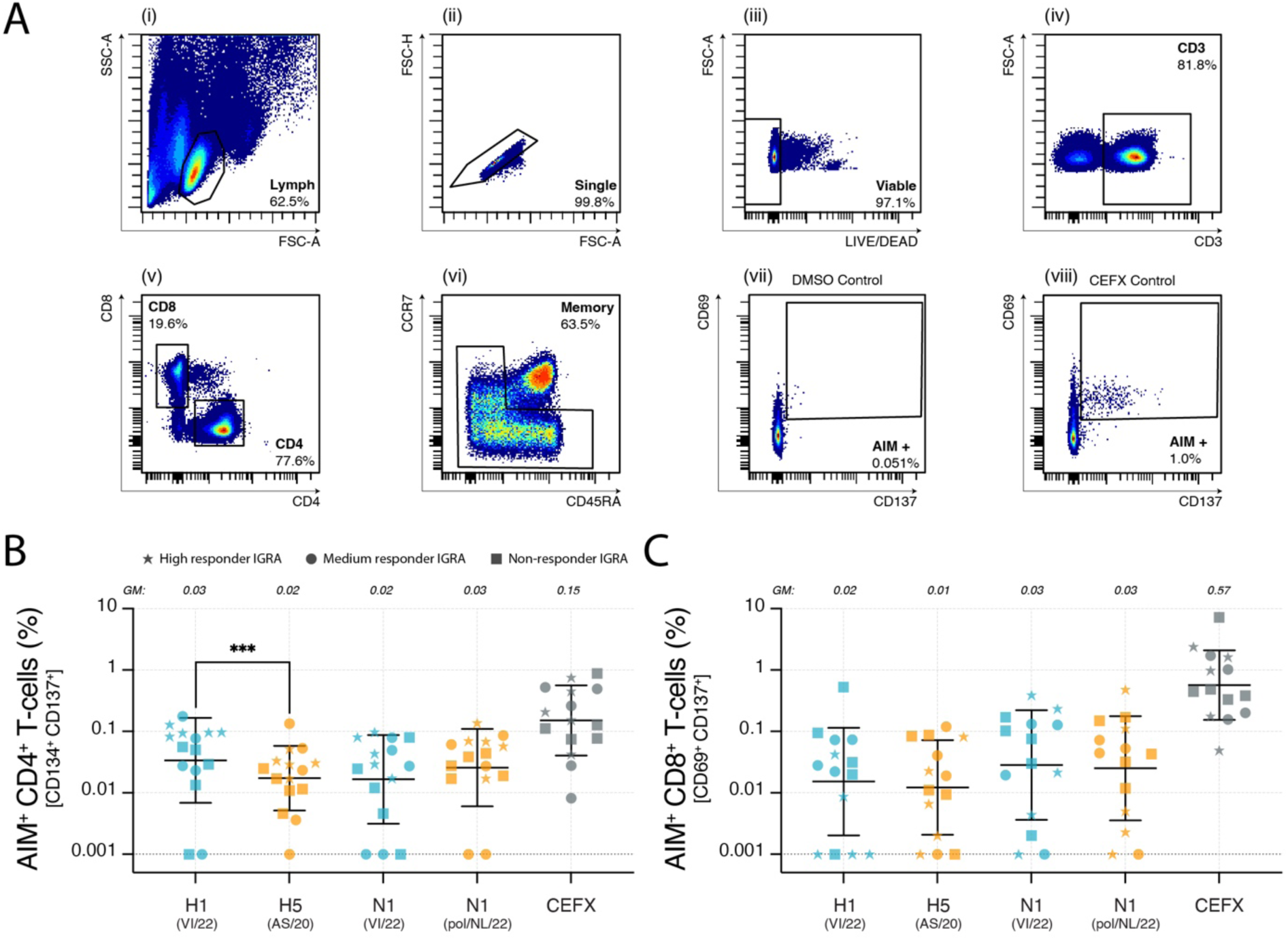
Detection of antigen-specific T-cells against HA and NA antigens from seasonal and A(H5) influenza viruses by AIM. **(A)** Gating strategy for AIM upregulation on virus-specific CD8^+^ T-cells: (i) lymphocytes were selected based on size and granularity, excluding (ii) doublets and (iii) dead cells, then (iv) CD3-expressing cells were selected, (v) dividing them into CD4^+^ and CD8^+^ subsets, and (vi) gating memory subsets (effector memory: CD45RA^-^CCR7^-^, central memory: CD45RA^-^CCR7^+^, and terminally differentiated effectors: CD45RA^+^, CCR7^-^). The activation of T-cells is measured as the percentage of CD134^+^/CD137^+^ or CD69^+^/CD137^+^ double-positive cells for CD4^+^ and CD8^+^ T-cells, respectively. For each donor, a DMSO-stimulated condition is included as a background control used for correction (vii, shown only for CD69^+^/CD137^+^ as an example). For each donor, a CEFX positive control is included to demonstrate the presence of activatable T-cells (viii, shown only for CD69^+^/CD137^+^ as an example). **(B and C)** Antigen-specific activation of (B) CD4^+^ and (C) CD8^+^ T-cells upon stimulation with A(H1) VI/22, A(H5) AS/20, N1 VI/22, or N1 pol/NL/22 peptide pools. AIM^+^ percentage was corrected for background DMSO values; the limit of detection is indicated at 0.001% AIM^+^. Bars indicate the GM with a 95% confidence interval. GM values for each peptide pool are listed at the top of each plot. Differences between paired samples of either A(H1) VI/22 and A(H5) AS/20, or N1 VI/22 and N1 pol/NL/22 were evaluated using the Wilcoxon matched pairs signed rank test. NS, no significance; *, p < 0.05; **, p < 0.01; ***, p < 0.001; ****, and p < 0.0001. Each dot represents an individual sample measurement from a total of 15 samples: 5 high (star), 5 medium (circle), and 5 IGRA non-responders (square). Abbreviations: AIM, activation-induced marker; DMSO, dimethyl sulfoxide; GM, geometric mean. Virus strain abbreviations can be found in **Supplemental Table 3**.

## Discussion

Population immunity is a vital component in tools that assess the pandemic risk of A(H5) influenza viruses, for example, the WHO TIPRA.^37^ To date, the data on the presence of cross-reactive immunity to A(H5) influenza viruses are fragmented, typically examining either antibody or T-cell-mediated responses alone, which provides an incomplete overview of protective immunity. Furthermore, due to the current pandemic threat from emerging A(H5) clade 2.3.4.4b influenza viruses, evaluations of population-based cross-reactive immune responses targeting these viruses need to be updated. Here, we examined the presence of both antibody and T-cell responses targeting novel A(H5) clade 2.3.4.4b influenza viruses in samples obtained from 107 HCW in the Netherlands in August and September 2024. We observed limited cross-reactive antibodies targeting the variable A(H5) HA head, containing the receptor binding domain, aligning with absent hemagglutination-inhibition activity. Nevertheless, we detected abundant cross-reactive antibodies to A(H5) HA0 antigens, likely directed at the conserved HA stalk, which mediated Fc-dependent ADCC. We detected an almost ubiquitous presence of NI antibodies against N1s from A(H5N1) viruses and T-cell responses against both HAs and NAs of avian influenza viruses. Importantly, these antibody and T-cell responses strongly correlated with responses against seasonal A(H1N1) influenza virus antigens, suggesting cross-reactive immunity induced by previous exposure.

The age-stratified analysis revealed that older individuals have higher binding IgG titers to conserved A(H5) HA0 antigens in the absence of antibodies reactive with the variable HA1, especially individuals over 60 years old. This indicates that these antibodies are likely reactive with the HA-stalk, and that the higher titers reflect the effect of repeated influenza virus exposure and continuous boosting of conserved epitopes. In fact, these findings confirm what was observed in recent studies, which indicated that non-neutralizing A(H5)-reactive antibodies targeting both historical and contemporary A(H5) influenza viruses are especially prevalent among older adults.^22,38,39^ Our correlation analyses showed that cross-reactive binding antibodies are likely induced by prior exposure to similar group 1 HA influenza virus subtypes (A(H1) or A(H2)). In both this study and current literature, instances where these cross-reactive binding antibodies are functional through HI or neutralization appear to be extremely rare.^38^

In the absence of direct HI activity, we examined other antibody functionalities. Nearly all HCW had detectable NA-inhibiting antibodies against seasonal influenza A viruses, which cross-inhibited NAs belonging to A(H5N1) influenza viruses, including a clade 2.3.4.4b virus, in accordance with a prior published study.^21^ In addition, we detected strong ADCC-mediating antibodies targeting the A(H5) clade 2.3.4.4b influenza AS/20 antigen, consistent with reports demonstrating that exposure to seasonal influenza viruses can induce broadly cross-reactive ADCC against older A(H5) influenza viruses.^26,27^ Notably, the ADCC activity in our cohort likely results from antibodies targeting the conserved HA-stalk, since it was observed in the absence of detectable HI A(H5) antibodies, which are typically directed toward the HA head.

In addition to functional antibodies, T-cells targeting avian influenza virus HAs and NAs were observed in the majority of HCW using IGRA. This recognition is likely attributed to cross-reactivity, due to the presence of conserved epitopes in seasonal and avian influenza viruses of the A(H5) clade 2.3.4.4b subtype, as previously demonstrated both computationally and experimentally.^39^ Therefore, individuals who have only been exposed to seasonal influenza A viruses likely possess cross-reactive T-cell memory that can recognize A(H5N1) viruses. AIM assays confirmed the presence of A(H5)-reactive T-cells; however, the percentages of antigen-specific cells and cytokine secretion in the supernatant were lower after A(H5) stimulation compared to A(H1) stimulation, indicative of cross-reactivity. However, the T-cell response was diverse, with the detection of both CD4^+^ and CD8^+^ T-cells, and balanced Th1/Th2 cytokine profiles.

Our findings show that, although HI antibodies targeting A(H5) influenza viruses were largely absent in the general population, robust functional ADCC and NA-inhibiting antibody responses to A(H5N1) clade 2.3.4.4b influenza viruses, elicited by prior exposure to seasonal influenza A viruses, were detected. ADCC-mediating antibodies have been described as a potential correlate of protection in humans, correlating with lower viral titers.^40^ NA-specific antibodies have been described to be a correlate of protection in both observational studies,^41,42^ clinical trials,^43^ and controlled human infection models.^44^ Whether these cross-reactive ADCC-mediating and NA-specific antibodies can mediate heterosubtypic immunity to A(H5) influenza viruses is supported by evidence from animal models. The presence of HA-cross-reactive ADCC-mediating antibodies correlated with reduced viral shedding upon A(H5N1) infection in macaques,^45^ whereas N1-inhibiting antibodies induced by exposure to seasonal influenza viruses provided protection upon A(H5N1) challenge of mice and ferrets.^46–48^ Specifically, for A(H5N1) clade 2.3.4.4b influenza viruses, heterosubtypic protection induced by exposure to A(H1N1) influenza virus has also been described in animal models, although the underlying correlates of cross-protection have not been completely elucidated.^49–51^ The protective role of T-cells has been studied less, and data for A(H5) clade 2.3.4.4b influenza viruses are lacking. However, heterosubtypic protection mediated by T-cells induced upon seasonal influenza A virus exposure against older A(H5) influenza viruses has been demonstrated in animal models.^52,53^ In humans, survivors of an A(H5N1) influenza virus infection developed strong polyfunctional T-cell responses, similar to the balanced cytokine profiles we observed in response to cross-reactive responses, implying that cell-mediated immunity can help to clear the virus and aid in recovery.^30^

Collectively, our findings have important public health implications. We demonstrate the widespread presence of complex, cross-reactive immune responses targeting novel A(H5) clade 2.3.4.4b influenza viruses in individuals unlikely to have been exposed to avian influenza viruses. While the protective value of these immune components in humans with complex immune backgrounds remains speculative, evidence from model systems suggests they can confer protection, potentially reducing disease severity and transmission. Disease outcome likely reflects a balance between inflammatory and virus-specific immune responses, which could also contribute to immunopathogenesis. Notably, we observed higher binding antibody responses to the A(H5) HA0 antigen in older individuals; however, key age groups, such as children, adolescents, and the elderly, were not represented in our cohort, precluding comprehensive age-stratified analyses. Other recent studies have reported age-related differences in immune responses to A(H5) influenza viruses, underscoring the potential value of targeting younger individuals in future vaccination efforts.

Importantly, our study highlights the need to expand the evaluation of cross-reactive immunity to include Fc-mediated antibody functions and T-cell responses, in addition to HI and neutralization assays. These immune components may significantly contribute to protection, especially when conventional correlates are absent. These findings support the concept that prior exposure to seasonal influenza viruses, through either infection, vaccination, or both, shapes a cross-reactive immune repertoire capable of recognizing conserved epitopes in A(H5) influenza viruses, thus offering broader protective potential at the population level.

## Data Availability

All data produced in the present work are contained in the manuscript.

## Acknowledgements

We would like to thank the Erasmus MC Virorunners for the SENTINEL study logistics, including but not limited to sampling, registration, and planning. We acknowledge the Erasmus MC department of Viroscience virus culture group for SENTINEL sample preparation. We thank the Erasmus MC Travel Clinic, and Simone Goeijenbier in particular, for logistical support in the SENTINEL study. We thank the members of the Dutch Vogelgriepconsortium for support, in particular Matthijn de Boer and Dirk Eggink. The study was funded by the Netherlands Organization for Health Research and Development (ZonMw, NCOH Pandemic Preparedness Research Kickstarter, grant agreement 10710022210003), European Union’s EU4Health program DURABLE (grant number 101102733), and the Dutch Ministries of Agriculture, Fisheries, Food Security and Nature and Health, Welfare and Sport. This project has been additionally funded with Federal funds from the National Institute of Health - National Institute of Allergies and Infectious Diseases (NIH-NIAID) contract no.75N93024C00056 and 75N93021C00014. The funding sources had no role in study design, data collection, analysis, interpretation of the data, or the decision to submit the paper for publication.

## Author contributions

Conceptualization: WS, SB, MPGK, CHGvK, RS, MR, RDdV. Data curation: WS, NHT, SB, CHGvK, RS, MR, RDdV. Formal analysis: MAP, WFR, AvdL, FC, RS, MR, RDdV. Funding acquisition: AS, MPGK, CHGvK, RS, MR, RDdV. Investigation: MAP, WFR, WS, LG, AvdL, FC, FV, TMB, NHT, SB. Methodology: MAP, WFR, LG, AvdL, FC, FV, TMB, GPvN, RS, MR, RDdV. Project administration: MAP, WFR, WS, BV, NHT, SB, CHGvK, RS, MR, RDdV. Resources: MAP, WFR, AG, GPvN, AS, RS, MR, RDdV. Supervision: CHGvK, RS, MR, RDdV. Visualization: MAP, WFR, MR, RDdV. Writing – original draft: MAP, WFR, MR, RDdV. Writing – review & editing: all co-authors.

