## Supplemental methods and figures for "Seasonal Influenza Exposure Elicits Functional Antibody and T-cell Responses to A(H5) Influenza Viruses in Humans"

**Study design and participants.** The SENTINEL study is an ongoing prospective follow-up study of HCW at Erasmus University Medical Center (Rotterdam, the Netherlands) to establish a biobank in which incidence, infection kinetics, and virus- and vaccine-specific immune responses for respiratory virus infections can be investigated. HCW over 18 years were invited to join the SENTINEL study. Sex or gender were not considered in the study design; the cohort comprises 73% female individuals (**Table 1**), reflecting the workforce of Erasmus University Medical Center. Vaccination history was collected through questionnaires.

**PBMC isolation.** Blood was diluted 1:1 in phosphate-buffered saline (PBS), layered on a density gradient (Leucosep, Greiner Bio-one), and PBMC were separated by centrifugation at 1,000 x g for 15 minutes. PBMC were washed twice in PBS and frozen in serum-free cryopreservation media (Bambanker, Nippon Genetics) in liquid nitrogen until use.

**Cells.** Human epithelial kidney 293T cells (ATCC-CRL-3216) were cultured in Dulbecco modified Eagle's medium (DMEM), high glucose (4.5 g/l) (Capricorn Scientific) supplemented with 10% foetal bovine serum (FCS; Sigma-Aldrich), 1 mM sodium pyruvate (Gibco), 1X non-essential amino acids (NEAA, Capricorn Scientific), 100 IU/mL penicillin (pen, Capricorn Scientific), 100 µg/mL streptomycin (strep, Capricorn Scientific) and 2 mM L-glutamine (L-glu, Capricorn Scientific). During basal cell culture, 293T cells were supplemented with 500 mg/mL geneticin (Gibco). Madin–Darby canine kidney (MDCK) cells (ATCC-CRL-2935) and humanized MDCK cells (hCK) were cultured in Eagle Minimum Essential Medium (EMEM) with Earle's balanced salt solution (Capricorn Scientific) supplemented with 10% FCS, 100 IU/mL pen, 100 mg/mL strep, 2 mM L-Glu, 10 mM 4-(2-hydroxyethyl)piperazine-1-ethane-sulfonic acid (HEPES, Capricorn Scientific), 1.5 mg/mL sodium bicarbonate (NaHCO3, Gibco), and 1X NEAA.[1] hCK cells were cultured with an additional 10 µg/mL blasticidin (InvivoGen) and 2 µg/mL puromycin (InvivoGen). NK92.05 cells, genetically modified to express a high-affinity CD16 Fc receptor (NK92.05-CD16), were kindly provided by Dr. K. S Campbell (Fox Chase Cancer Center). NK92.05 cells were cultured in Alpha-MEM (Capricorn Scientific) supplemented with 2.2 g/L sodium bicarbonate (NaHCO3, Gibco), 0.0001 M 2-mercaptoethanol (BME, Sigma-aldrich), 200 mM L-glu (Capricorn Scientific), 0.2 mM myo-inositol (Sigma-aldrich), 10% horse serum (Sigma-Aldrich), 10% FCS (Sigma-Aldrich), 0.004 mM folic acid (Sigma-Aldrich), 1 mM sodium pyruvate (Sigma-Aldrich), 100 IU/mL pen (Capricorn), 100 µg/mL strep ( Capricorn Scientific), additionally supplemented with 100 IU/mL recombinant human IL-2 (Proleukin rIL-2, kindly provided by the Erasmus MC Hospital Pharmacy) two times a week. Cells were cultured at 37°C and 5% $CO_2$.

**Protein microarray.** A protein microarray (PMA) assay was performed to measure the presence of serum binding IgG antibodies directed to a large panel of commercially acquired HA recombinant proteins from the A(H1), A(H2), A(H3), A(H5), A(H7), and B subtypes as described previously.[2] Antigens representing both the HA1 (containing the globular head and a small proportion of the stem) and the HA0 were used (**Supplemental Table 2** for details on origin, conformation, and supplier). Briefly, recombinant proteins were printed using a non-contact array spotter (sciFLEXARRAYER SX, Scienion) on 24-pad coated (Oncyte Avid) glass slides (Grace Bio-Labs). Optimal antigen concentrations were determined for each antigen through checkerboard titrations.[3] To prevent non-specific binding to the nitrocellulose surface, dried slides were incubated with Blocker™ Blotto buffer (Thermo Fisher Scientific Inc.) for one hour at 37°C in a moist chamber. Between each step, slides were washed three times using a protein array wash buffer (Whatman). Sera were pre-diluted in Blocker™ Blotto blocking buffer in Tris-buffered saline (TBS) containing 0.1% Surfact-Amps 20 (Thermo Fisher Scientific Inc.), and then four-fold serially diluted. The diluted sera were transferred to the slides and incubated for one hour at 37°C in a moist chamber. After washing, an AF647-conjugated goat-anti-human IgG (Fc-fragment specific, Jackson ImmunoResearch) was added and allowed to bind for one hour at 37°C in a moist chamber. After final washing steps, fluorescence median signals were measured using the microarray scanner InnoScan 910 AL (Innopsys) and analysed using mapix software. Titers (range 20-20,480) were determined as the serum concentration that induced a 50% response in the concentration-response curve. Samples obtained post A(H5) vaccination from a previous study conducted at the Erasmus Medical University Center were included as controls for the A(H5) antigens (validation is shown in **Supplemental Figure 7**).[4] For graphing and geometric mean titer (GMT) calculations, threshold titers of <20 were set to a value of 10 to indicate undetectable values.

**Production of plasmids.** To produce plasmids for recombinant virus production, viral RNA was extracted from in-house virus isolate stocks using the High-Pure RNA isolation Kit (Roche), following the manufacturer's protocol. Isolated RNA was subsequently utilized as a template for viral complementary DNA (cDNA) production and amplification of individual gene segments by reverse transcription PCR (RT-PCR) using the SuperScript™ III One-Step RT-PCR System with Platinum™ Taq High Fidelity DNA Polymerase (Invitrogen), following the manufacturer's instructions. Segment PCR primer sets included 5'-GGGGGGAGCAAAAGCAGGGG-3' and 5'-CCGGGTTATTAGTAGAAACAAGGGTGTT-3' for HA, and 5'-GGGGGGAGCAAAAGCAGGAGT-3' and 5'-CCGGGTTATTAGTAGAAACAAGGAGTT-3' for NA. The HA or NA gene segments were cloned into a previously described modified version of the pHW2000 plasmid by seamless cloning using the GeneArt™ Seamless Cloning kit (Thermo Fisher Scientific Inc.) or the In-Fusion® HD Cloning Kit (Takara Bio),[5] according to the manufacturer's protocol. If applicable, the multibasic cleavage site was removed by site directed mutagenesis using specific primers. When the required viruses were not available in-house, synthetic genes comprising either HA or NA sequences were synthesized by Proteogenix. HA sequences were ordered with a monobasic cleavage site when applicable. The recommended viruses for inclusion in the trivalent or quadrivalent influenza virus vaccine for the 2024-2025 northern hemisphere (e.g., A/Victoria/4897/2022 [VI/22], A/Thailand/8/2022 [TH/22], B/Austria/1359417/2021 [AU/21] and B/Phuket/3073/2013 [PH/13]), as well as the avian influenza A/Anhui/1/2005 [AN/05], A/European Polecat/Netherlands/1/2022 [pol/NL/22], A/Hunan/09285/2021 [HU/21], and A/Shearwater/Western Australia/2576/1979 [shear/WA/79], strains were present in-house as virus isolates. VI/22, TH/22, PH/13, and AN/05 were ordered from the National Institute for Biological Standards and Control (NIBSC 22/316, 23/204, 21/132, and 07/290, respectively), AU/21 was passaged once in MDCK-SIAT1 (cells stably transfected with cDNA of human α-2,6-sialyltransferase) and three time on MDCK cells and kindly provided by Ms. A. Rattigan (Francis Crick Institute). HU/21 was kindly provided by the Chinese Center of Disease Control and Prevention[6] and shear/WA/79 by Dr. R.G. Webster (St. Jude Children's Research Hospital).[7] Pol/NL/21 was isolated from a carcass of a naturally infected European polecat (*Mustela putorius*) as part of a citizen reporting system of dead wildlife to the Dutch Wildlife Health Centre (DWHC).[8] The avian influenza A/Astrakhan/3212/2020 (AS/20) gene segment was synthetically ordered. Accession numbers can be found in the supplementary information (**Supplemental Table 4**).

**Production and propagation of recombinant viruses and virus isolates.** Recombinant influenza viruses were produced by reverse genetics using the eight-plasmid system, as described previously.[9] For the HI assays (described below), seasonal influenza viruses were virus isolates, and A(H5) influenza viruses were recombinant viruses carrying the HA of interest, without the multibasic cleavage site if applicable, with the seven remaining gene segments of the attenuated strain A/Puerto Rico/8/1934 (PR/8) high yield (HY) (**Supplemental Table 4**).[9] For the ELLA and NI assays, recombinant viruses contained the A(H15) shear/WA/79 and the NA of interest, in combination with the remaining six gene segments of PR/8 HY (**Supplemental Table 4**). Recombinant viruses were generated under biosafety level 2 conditions. Viruses were amplified in MDCK cells, and viral presence was assessed by a hemagglutination assay with a final concentration of 0.5% turkey red blood cells in PBS. The seasonal influenza viruses VI/22, TH/22 and PH/13 were propagated in eggs, while AU/21 was amplified in hCK cells. Sequences of HA and NA genes from viruses of interest for a particular assay were confirmed using the BigDye™ Terminator v3.1 Cycle Sequencing Kit (Applied Biosystems) and the 3500xL Genetic Analyzer (Applied Biosystems).

**Hemagglutination inhibition (HI) assay.** HI assays were performed to evaluate the presence of functional HA-specific serum antibodies. As seasonal influenza viruses, the strains included in the 2024-2025 northern hemisphere quadrivalent influenza vaccine were selected: VI/22, TH/22, AU/21, and PH/13.[10] As A(H5) influenza viruses two World Health Organization (WHO) candidate vaccine viruses (CVVs, AN/05 (H5 clade 2.3.4) and AS/20 (H5 clade 2.3.4.4b)) and a mammalian virus from 2022 pol/NL/22 (H5 clade 2.3.4.4b) were selected.[8] Recombinant viruses in PR/8 HY background and virus isolates (**Supplemental Table 4**) were used for HI assays using either horse or turkey red blood cells (hRBCs or tRBCs, respectively). hRBCs were used to test for the presence of functional antibodies targeting avian influenza A viruses, whereas tRBCs were used for human influenza A and B viruses. Using hRBCs instead of the more commonly employed tRBCs has been shown to enhance assay sensitivity when testing human serum with avian influenza A viruses.[11] Ferret sera from animals infected with homologous or antigenically related viruses were included as a positive control for A(H5) influenza viruses.

*HI assay with horse red blood cells.* hRBCs were washed three times with PBS by centrifugation at 750 x g for 10 minutes at room temperature. Solutions of 2% and 10% hRBCs in PBS were prepared and stored at 4°C for a maximum of one week. The HI assay was always performed at least one day after washing to ensure optimal RBC settlement. To prevent aspecific agglutination, sera were absorbed with an equal volume of 10% hRBCs, and the serum was incubated for one hour at 4°C with careful mixing every 20 minutes. Sera were then treated overnight at 37°C in a 1:6 ratio with in-house produced *Vibrio cholerae* filtrate, which contained receptor-destroying enzyme (RDE), followed by heat inactivation at 56°C for 1 hour to avoid aspecific inhibition. Sera were further diluted with 0.5% bovine serum albumin (BSA, Sigma) in PBS (0.5% BSA-PBS) to a final 1:20 dilution in 50 µL and subsequently two-fold serially diluted in V-bottom microtiter plates. To each well, 25 µL PBS comprising 4 hemagglutinating units (HAU) of recombinant virus was dispensed. Plates were mixed by tapping and then incubated at 37°C for 30 minutes. After this, 25 µL of 2% hRBCs was added to all wells. Plates were individually tapped, and HI titers were read after 1.5 hours of incubation at 4°C. The HI titer was expressed as the inverse of the highest serum dilution where hRBC agglutination was completely inhibited. For each serum, a control containing 25 µL of PBS instead of 4 HAU recombinant virus was included as a negative control. If background agglutination was observed, HI assays with the corresponding sera were repeated, in which the sera were absorbed twice instead of once. The lowest detectable HI titer was 10, corresponding to partial agglutination in the 1:20 serum dilution condition. For graphing and GMT calculations, threshold titers of <10 were set to a value of 5 to indicate undetectable values.

*HI assay with turkey erythrocytes.* In-house obtained tRBCs were stored at 4°C for a maximum of one week. Washing of tRBCs was performed by centrifuging three times at 205 x g for 10 minutes at room temperature with PBS. Solutions of 1% and 10% tRBCs in PBS were prepared. One volume of serum was pre-treated with five volumes of in-house manufactured *Vibrio cholerae* filtrate containing RDE overnight at 37°C. The following day, RDE was heat-inactivated at 56°C for 1 hour. Afterward, sera were incubated with an equal volume of 10% tRBCs at 4°C for 1 hour, with careful mixing every 30 minutes. The HI assay was further processed as described above with the following alterations: 0.5% BSA-PBS was substituted by PBS, the assay was executed in U-bottom plates with 25 µL of 1% tRBCs, and HI patterns were read after 1 hour of incubation at 4°C. In separate HI assay runs, a selected subset of serum samples and antigens were incorporated to enable cross-validation and assess reproducibility between experiments. HI titers showed an average 1.8-fold difference between runs, with a range of 1- to 8-fold, indicating acceptable inter-assay variability.

**Neuraminidase inhibition (NI) assay.** The enzyme-linked lectin assay (ELLA) and neuraminidase inhibition (NI-ELLA) assay were performed to determine the presence of functional NA-specific serum antibodies against a panel of selected antigens (**Supplemental Table 4** for details on the viruses used for these assays).[11] Recombinant A(H15) shear/WA/79 viruses with the NA of interest in the PR8 HY backbone were generated and used to mismatch the HA and thereby prevent the interference of HA-specific antibodies.

*ELLA.* Recombinant viruses were titrated to measure the enzymatic sialidase activity of NA by detecting NA-induced desialylation of highly glycosylated proteins to determine the optimal virus dilution that resulted in NA activity within the linear range of the titration curve (e.g., 50% of the total NA activity). To this end, Nunc-Immuno™ MicroWell™ 96-well solid plates (Thermo Fisher Scientific Inc.) were coated with 25 µg/mL fetuin (100 µl/well, Sigma) in 1X KPL coating solution concentrate (SeraCare) and stored at 4°C for at least 24 hours and a maximum of 2 months. Two-fold serial dilutions of recombinant viruses (antigen) in sample diluent (consisting of 1X Dulbecco's PBS with 0.33 g/L calcium chloride ($CaCl_2$) and 0.1 g/L magnesium chloride ($MgCl_2$, DPBS, Gibco) supplemented with 1% BSA and 0.5% Tween® 20 (Sigma-Aldrich)) were made. Fetuin-coated plates were washed three times with PBS-Tween® 20 (PBST) and

50 µL of antigen serial dilutions were transferred in duplicate to fetuin-coated plates containing 50 µL sample diluent. On each plate, no antigen was added to the last column. Plates were incubated at 37°C, 5% $CO_2$ for 16 hours. The following day, plates were washed six times with PBST and incubated with 100 µL/well of 1 µg/mL peanut agglutinin conjugated to horseradish peroxidase (PNA-HRPO, Sigma-Aldrich) in conjugate diluent (DPBS-1% BSA) at room temperature for two hours in the dark. After incubation, the plates were washed three times with PBST and developed in the dark with 100 µL/well of freshly prepared *o*-Phenylenediamine dihydrochloride (OPD, Sigma-Aldrich) substrate according to the manufacturer's instructions. The reaction was stopped after exactly 10 minutes with 100 µL/well of 1N sulfuric acid ($H_2SO_4$). The optical density (OD) of each plate was immediately measured at 490 nm on a Tecan plate reader (Infinite 200 96-well plate reader, Tecan). NA activity was determined by subtracting the mean background of each plate (average from wells in which no antigen was added) from the absorbance of wells with antigen. The optimal antigen dilution for the NI-ELLA was determined as 50% of the total NA activity.

*NI-ELLA.* To prevent aspecific inhibition, one volume of serum was incubated overnight at 37°C with five volumes of 10-fold diluted in-house produced RDE in sample diluent, followed by an eight-hour RDE inactivation at 56°C. Treated serum samples were two-fold serially diluted in sample diluent, starting at a 1:10 dilution. Fifty µL of the serial dilutions was transferred in duplicate to fetuin-coated plates, which had been previously washed three times with PBST and contained 50 µL of antigen diluted in sample diluent accordingly to the results of the ELLA assay. On each plate, serum was omitted from the last two columns, and antigen was additionally omitted from the final column. Plates were incubated at 37°C, 5% $CO_2$ for 16 hours. The rest of the assay was performed as described above for the ELLA. The NI titer was determined separately for each plate. First, the background signal, measured from the 8 wells lacking both antigen and serum (final column), was subtracted from the signals of all other wells. The mean maximal NA activity was then calculated using the 8 wells containing antigen but no serum (second-last column). For each serum dilution, NA activity was expressed as the percentage of the mean maximal NA activity. The NI titer was defined as the reciprocal of the highest serum dilution that resulted in at least 50% inhibition of NA activity. For graphing and geometric mean titer (GMT) calculations, threshold titers of <10 were set to a value of 5 to indicate undetectable values.

To enable a comparison across NI-ELLA assays performed on different days, overlapping serum samples and antigens were included in each experiment. NI titers from repeated conditions showed an average 1.4-fold difference, with a range of 1- to 4-fold, indicating acceptable inter-assay variability.

**Antibody-dependent cellular cytotoxicity.** The degranulation of natural killer (NK) cells through antibody Fc-mediated effector functions was evaluated utilizing the previously described antibody-dependent cellular cytotoxicity (ADCC) assay.[12] In summary, high-binding flat-bottom 96-well plates (Immunolon, Fisher Scientific) were coated with 100 ng/well of HEK293 cell-generated A(H1) VI/22 HA0 (Sino Biological) or baculovirus-insect cell-generated A(H5) AS/20 HA0 (**Supplemental Table 2**) at 4°C overnight. After coating, the plates were washed twice with 200 µL of PBS per well, then blocked with PBS supplemented with 5% BSA (Sigma-Aldrich) and incubated at 37°C and 5% $CO_2$ for 30 minutes. Afterward, the plates were washed 5 times with 200 µL of PBS per well and incubated with sera serially diluted four-fold in PBS, starting at a 1:40 dilution, at 37°C and 5% $CO_2$ for 2 hours. A PBS control and a standard reference pool composed of five sera obtained post A(H5) vaccination from a previous study exhibiting high HI and neutralization titers were included on every plate.[4] Following the incubation with serum, plates were washed, and 100,000 NK92.05-CD16 cells were added, supplemented with an anti-CD107a antibody (V450-conjugated, 1:100, clone H4A3, BD), Golgistop (0.67 µL/mL, BD), and GolgiPlug (1 µL/mL, BD). The plates were incubated at 37°C and 5% $CO_2$ for five hours, subsequently washed with PBS, and viable NK cells were stained with an anti-CD56 antibody (PE-conjugated, 1:25, clone B159, BD) and LIVE/DEAD Fixable Aqua Dead Cell Stain (1:100, AmCyan, Invitrogen) at 4°C for 30 minutes. Plates were fixed in Cytofix/Cytoperm (BD) at 4°C for an additional 30 minutes. NK92.05-CD16 cells were acquired using a FACSLyric (BD), and activated cells were identified as LIVE/CD56+/CD107a+, degranulating NK cells. S-curves were generated by using the lowest and highest percentages of NK92.05 cell degranulation normalized to the reference pool on each plate, from which a 30% endpoint was calculated. To correct for inter-assay variation across runs, a correction factor was applied to each sample by comparing its plate reference serum 30% endpoint titer to the average 30% endpoint titer of the A(H5) reference serum across all plates. A threshold titer of 40 was utilized in this assay.

**Peptide pools.** Overlapping peptide pools (15-mers with 10 amino acid overlap) that span the HAs and NAs of the following viruses were designed and produced in-house: HA pools covering seasonal A(H1), seasonal A(H3), seasonal B/Vic, seasonal B/Yam, A(H5) clade 2.3.4, and A(H5) clade 2.3.4.4b were generated. Additionally, NA pools covering seasonal N1, seasonal N2, avian N1, avian N6, and avian N8 were generated (**Supplemental Table 5** and **6** for details on the peptide pools used for these assays). Peptides were synthesized as crude material (TC peptide lab, San Diego), resuspended in dimethyl sulfoxide (DMSO), pooled, and sequentially lyophilized as previously described.[13] The overlapping peptide pool of measles virus N protein (15-mer with 10 amino acid overlap) was synthesized and lyophilized commercially (Biosynth). Commercially available overlapping peptide pools (JPT Peptide Technologies) spanning the SARS-CoV-2 spike (S) protein or a pool of known infectious agent epitopes (CEFX) were used as positive control.

**T-cell profiling through interferon gamma (IFN-γ) release assay (IGRA).** To determine T-cell response profiles, IFN-γ production was measured after stimulation of whole blood with peptide pools covering the HA or NA proteins of influenza A or B viruses of interest, the N protein of measles virus, the S protein of SARS-CoV-2 or CEFX. Peptide pools (1.0 µg/mL) were added to 200 µL of whole blood collected in lithium heparin tubes (BD) and incubated in 96-well plates at 37°C and 5% $CO_2$ for 20 hours. As a negative control, blood was stimulated with an equimolar amount of DMSO. As a positive control, blood was stimulated with an aspecific mitogen, phorbol 12-myristate 13-acetate (PMA) and ionomycin (Sigma-Aldrich). Following incubation, whole blood was centrifuged at 1,000 × g for 15 minutes. Next, plasma was collected and IFN-γ was measured using a commercial pre-coated anti-IFN-γ ELISA, which is part of the QuantiFERON SARS-CoV-2 ELISA kit (Qiagen). In short, 50 µL of plasma was incubated for 2 hours with the enzyme-conjugate solution (1:1). Pre-coated ELISA plates were washed 4 times with wash buffer, developed for 10 minutes using substrate solution, and the colour development was terminated using a stop solution, according to the manufacturer's instructions. ELISA plates OD was measured at a wavelength of 420 nm with a reference filter wavelength of 620 nm on a Tecan plate reader (Infinite 200 96-well plate reader, Tecan). IFN-γ levels were calculated from a standard curve with known IFN-γ concentrations provided with the QuantiFERON SARS-CoV-2 ELISA kit and depicted as IU/mL. All values below the detection limit of 0.01 IU/mL were set to that value.

**Activation-induced marker (AIM) T-cell assay.** The AIM assay was conducted to phenotype HA and NA-specific T-cells in a selection of IGRA high, medium, and non-responders. In summary, a quartile analysis was conducted on the IGRA results from A(H1) VI/22, A(H5) AS/20, N1 VI/22, and N1 pol/NL/22. Individuals who averaged in the 75th, 50th, or 25th percentile were classified as IGRA high, medium, or non-responders, respectively. Five individuals were then chosen from each subgroup for the AIM assay. After thawing, $1 \times 10^6$ PBMC were resuspended in RPMI 1640 Medium, with L-Glutamine, with 25 mM HEPES , which was supplemented with 10% human pooled serum (HPS, Sanquin), 100 IU/mL pen, and 100 IU/mL strep (R10H medium), and stimulated with peptide pools (1 µg/mL) covering the HA or NA proteins of influenza A or B viruses of interest. As a negative control, PBMC were stimulated with an equimolar amount of DMSO. As a positive control, PBMC were stimulated with a specific pool of immunodominant peptides (CEFX). Following stimulation, supernatants were stored at -80°C for further use for cytokine quantification. PBMC were stained for surface markers at 4°C for 30 minutes with the following antibodies conjugated with the indicated fluorophores at their respective dilutions: anti-CD3 (PerCP, Clone SK7, BD, 1:25), anti-CD4 (V450, Clone L200, BD, 1:50), anti-CD8 (FITC, Clone DK25, Dako, 1:25), anti-CD45RA (PE−Cy7, Clone L48, BD, 1:50), anti-CCR7 (BV711, Clone 150503, BD, 1:25), anti-CD69 (APC−H7, Clone FN50, BD, 1:50), anti-CD137 (PE, Clone 4B4-1, Miltenyi, 1:50), and anti-OX40 (CD134) (BV605, Clone L106, BD, 1:25). LIVE/DEAD™ Fixable Aqua Dead Cell staining was included (AmCyan, 1:100). Lymphocytes were gated based on forward and side scatter area. LIVE lymphocytes were gated, and T-cells were identified as CD3$^+$ cells and subdivided into CD4$^+$ or CD8$^+$ subsets. Memory subsets were classified as either CD45RA$^+$/CCR7$^+$ (naïve, TN), CD45RA$^-$/CCR7$^+$ (central memory, TCM), CD45RA$^-$/CCR7$^-$ (effector memory, TEM), or CD45RA$^+$/CCR7$^-$ (terminally differentiated effectors, TEMRA). Influenza HA or NA reactive T-cells were gated as activated T-cells (CD137$^+$/CD134$^+$ for CD4$^+$ T-cells, or CD137$^+$/CD69$^+$ for CD8$^+$ T-cells), following the exclusion of TN cells. The gating of subsets and activated cells was established based on the DMSO-stimulated sample on a per-donor basis. Samples containing fewer than 10,000 CD4$^+$ or 5,000 CD8$^+$ T-cells were excluded from analysis. AIM$^+$ values were adjusted for their donor-matched DMSO condition. An arbitrarily chosen lower limit of detection (LLOD) of 0.001% was set to reliably identify AIM$^+$ cells within the CD4$^+$ or CD8$^+$ T-cell gates.

**Legendplex.** Cytokine detection in AIM supernatants was performed to detect cytokines released in response to stimulation with HA and NA peptides. The stored supernatants were thawed and analysed using a T-helper (Th) cytokine panel kit (Legendplex, BioLegend). This panel comprised interleukin (IL)-2, IL-4, IL-5, IL-6, IL-9, IL-10, IL-13, IL-17A, IL-17F, IL-22, IFN-γ, and TNF-α. To identify Th1 T-cells, the levels of IL-2, IFN-γ, and TNF-α were measured. To detect Th2/Th9 T-cells, the levels of IL-4, IL-5, IL-9, IL10, and IL13 were measured. Cytokines IL-6, IL-17A, IL-17F, and IL-22 were excluded from downstream analysis due to their early (IL-6) or delayed secretion kinetics, which typically peak beyond the 24-hour stimulation window used in this assay. As a result, their levels at this time point were either low or inconsistent, which limited their interpretability. Samples were stained according to the manufacturer's instructions. In short, the supernatants were thawed on ice, centrifuged, and incubated with monoclonal capture antibody-coated beads for two hours under constant agitation. Following this, the beads were washed and incubated with biotin-labelled detection antibodies for one hour, and subsequently with streptavidin-PE for thirty minutes. After staining, the beads were acquired using a FACSLyric (BD). The acquired data were analysed with LEGENDplex V8.0 software (BioLegend). The quantity of each cytokine was determined based on the intensity of the streptavidin-PE signal and a freshly prepared standard curve.

The results were expressed as pg/mL after subtraction of the DMSO-stimulated control values. Sample values that were negative after DMSO subtraction were adjusted to an arbitrary value of 0.001 pg/mL.

**Statistical analysis.** Baseline characteristics of the HCW cohort were summarized as counts and percentages for categorical variables and as medians with interquartile ranges (IQRs) for continuous variables. Principal component analysis (PCA) was performed on all log10(x+1) transformed and z-scored PMA-measured HA0-binding IgG titers. PCA was conducted using the R packages FactoMineR (v2.11), ggplot2 (v3.4.4), and ellipse (v0.5.0) within RStudio (V4.4.0). The number of principal components explaining ~80% of the total variance was determined through screeplot analysis. K-means clustering was applied to the principal component scores using base R functions, with the nstart parameter set to 25 to ensure cluster stability. A 90% confidence ellipse was generated for the dominant cluster using its covariance matrix. Data normality was assessed by applying appropriate transformations, including $\log_2$ (for NI), log4 (for PMA and ADCC), or $\log_{10}$(x+1), followed by visual inspection with histograms and Q-Q plots, and statistical evaluation using the Shapiro–Wilk test. Analyses were performed in RStudio (v4.4.0) using ggplot2 and base R functions. Spearman correlation coefficients were calculated using GraphPad Prism (v10.5.0). Intra-individual differences in T-cell responses between dimethyl sulfoxide (DMSO) control and antigen-stimulated conditions were evaluated with the non-parametric Wilcoxon signed-rank test, followed by the Benjamini-Hochberg procedure in RStudio (v4.4.0). $AIM^+$ and cytokine T-cell responses were evaluated with the Wilcoxon matched-pairs signed rank test. Age-stratified heatmaps of binding IgG responses to HA1 and HA0 antigens were generated using the ComplexHeatmap package (v2.18.0), along with dplyr (v1.1.4) and circlize (v0.4.16), following $\log_{10}$(x+1) transformation and per-antigen z-score normalization; z-scores were Winsorized to the 5th and 95th percentiles to minimize the impact of outliers. Differences in A(H5) HA0 binding IgG antibody responses across age groups were assessed using the Kruskal-Wallis test, followed by Dunn's multiple comparisons test. Correlation matrices were generated using Spearman correlation coefficients from $\log_{10}$(x+1) transformed data without additional normalization. P values < 0.05 were considered statistically significant.

### Supplemental Figures

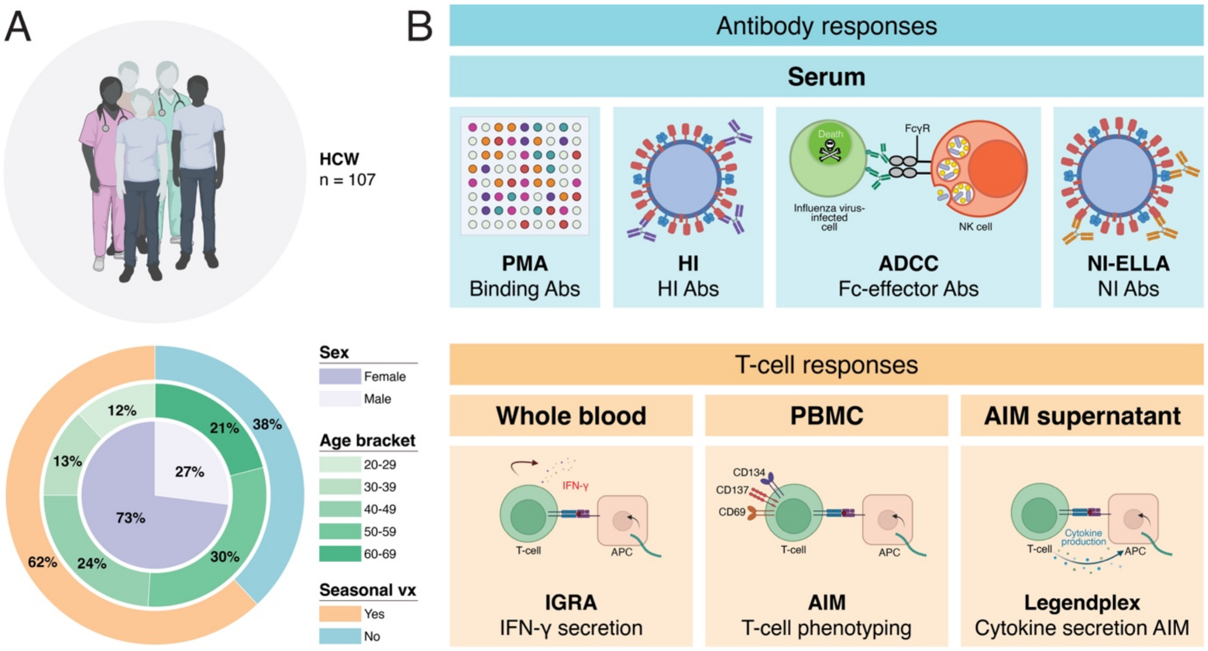


**Supplemental Figure 1. Cohort characteristics and schematic overview of the serological and T-cell assays. (A)** Schematic overview of cohort inclusions (n=107 healthcare workers, top panel) and cohort characteristics (biological sex, age bracket, and history of prior seasonal vaccination, bottom panel). Characteristics are summarized in **Table 1**. **(B)** Schematic overview of serological assays (top panel) and T-cell assays (bottom panel) performed in the scope of this study. Abbreviations: HCW, healthcare workers; vx, vaccination; PMA, protein microarray assay; HI, hemagglutination inhibition; ADCC, antibody-dependent cellular cytotoxicity; NI, neuraminidase inhibition; ELLA, enzyme-linked lectin assay; PBMC, peripheral blood mononuclear cells; AIM, activation-induced marker; IGRA, IFN-γ release assay.

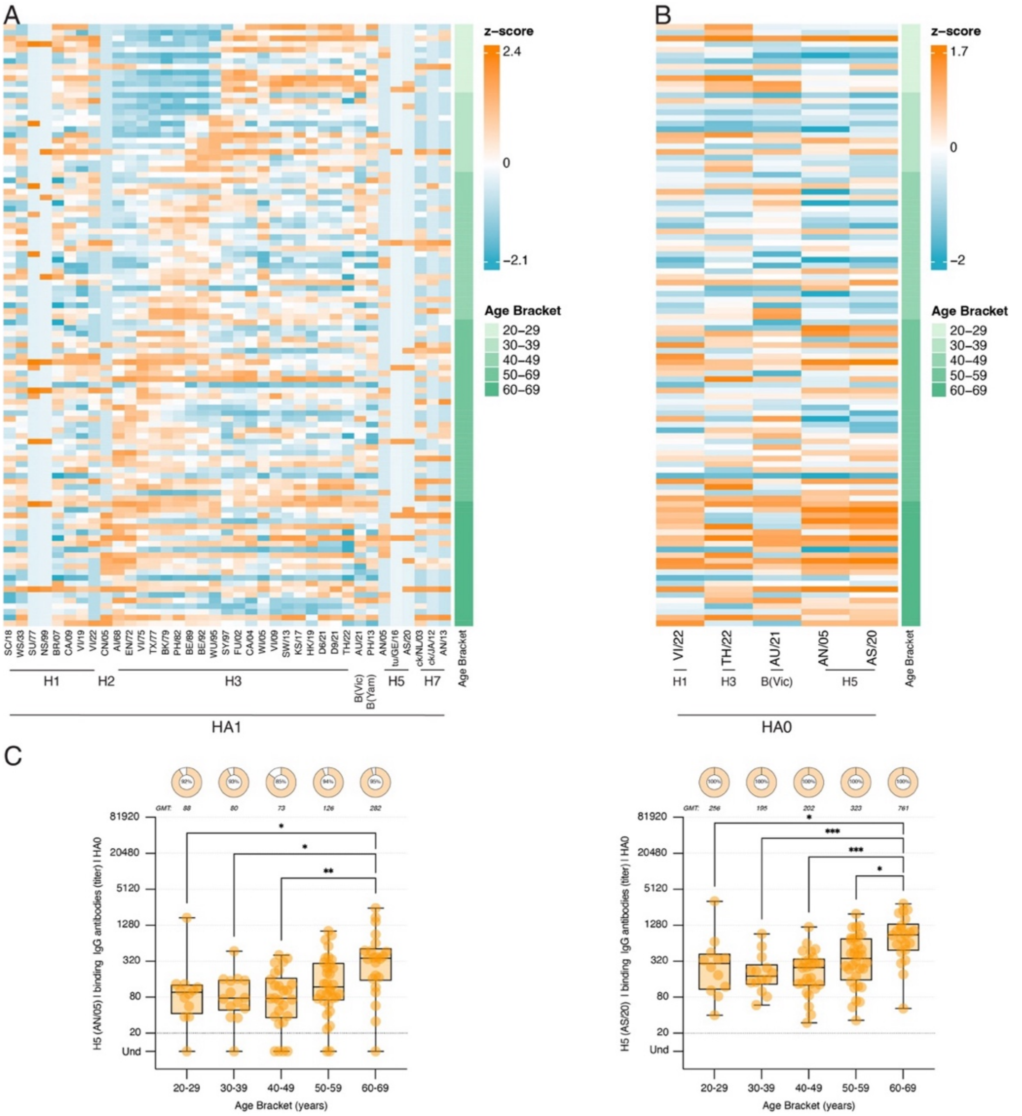


**Supplemental Figure 2. Binding serum IgG antibodies measured by PMA, stratified by age group**. PMA titers against various HA1 and HA0 antigens, as shown in **Figure 1,** are visualized as **(A** and **B)** heatmaps stratified by age group or **(C)** individual antigen box plots. **(A)** Binding IgG antibody levels targeting the HA1 subunit of a panel of seasonal (A(H1), A(H2), A(H3), B(Vic), and B(Yam)) and avian (A(H5), A(H7)) influenza viruses. **(B)** Binding IgG antibody levels targeting the HA0 of the 2024-2025 northern hemisphere seasonal influenza virus vaccine antigens (A(H1), A(H3), B(Vic), and B(Yam)) and A(H5) influenza viruses. Values were $\log_{10}(x+1)$ transformed, then centred, and standardized (z-scored) for each antigen. To minimize the impact of outliers, z-scored values were Winsorized to the 5th and 95th percentiles. Warmer colours indicate higher z-scores, while cooler colours show lower z-scores. Each row represents an individual serum sample (ordered from youngest to oldest). Each column corresponds to one antigen. **(C)** Box plots depict binding IgG antibodies to HA0 of A(H5) clade 2.3.4 AN/05 and A(H5) clade 2.3.4.4b AS/20 for pre-defined age brackets. The central box spans the IQR, with the horizontal line indicating the median. Whiskers extend to the minimum and maximum values. The dashed horizontal line represents the threshold titer of 20. Samples with an unmeasurable PMA titer are indicated as undetectable. Each dot represents an individual sample measurement from a total of 106 samples. Differences between the age brackets were assessed using the Kruskal–Wallis test, followed by Dunn's multiple comparisons test. P-values were adjusted for multiple comparisons. NS, no significance; *, $p < 0.05$; **, $p < 0.01$; ***, $p < 0.001$; ****, and $p < 0.0001$. Heatmaps were generated in R (version 4.4.0) using packages dplyr (version 1.1.4), ComplexHeatmap (version 2.18.0), and circlize (version 0.4.16). Abbreviations: PMA, protein microarray assay; ; IQR, interquartile range. Virus strain abbreviations can be found in **Supplemental Table 3**.

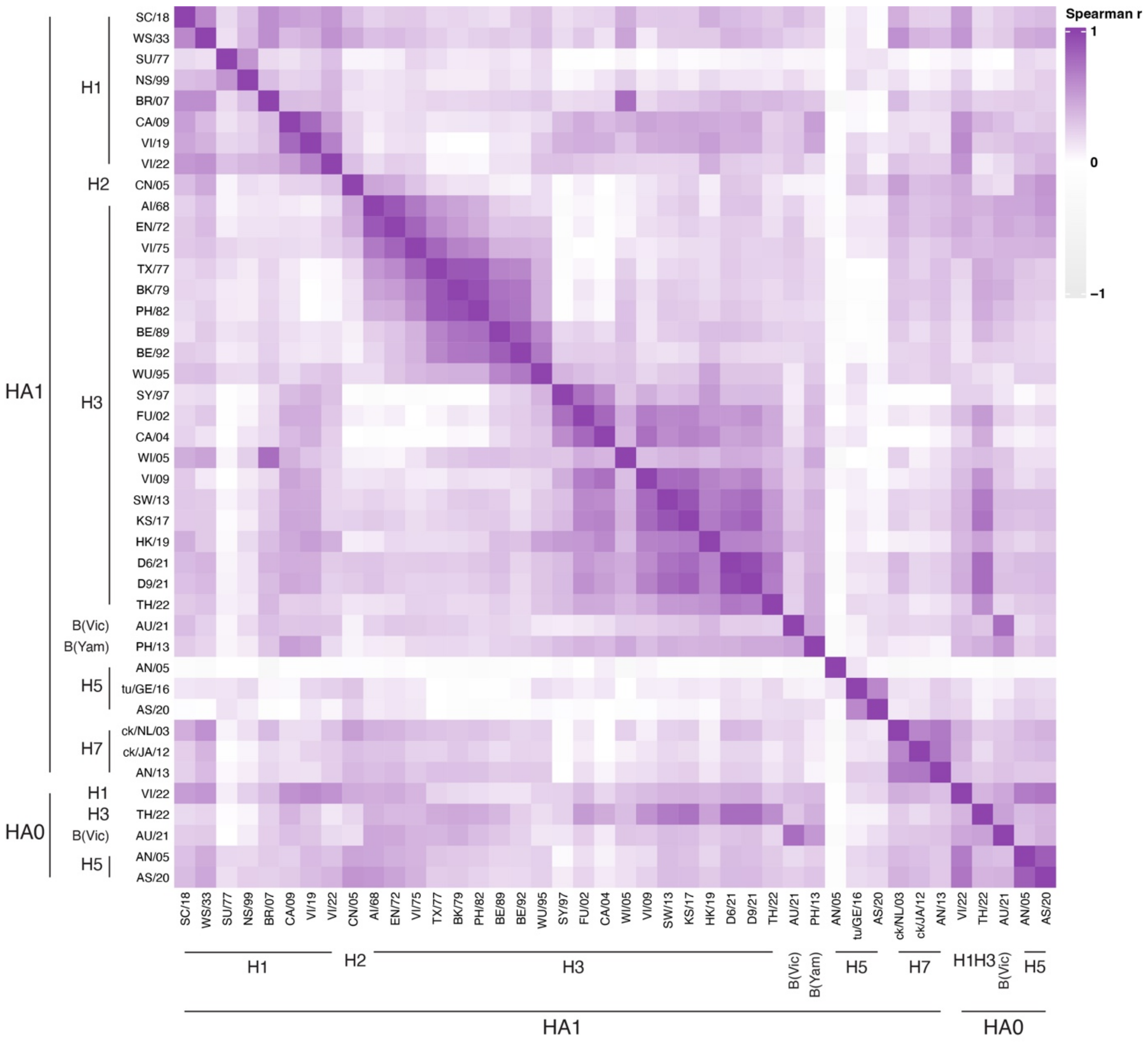


**Supplemental Figure 3. Spearman correlation matrix of HA0 and HA1 antigen-specific responses.** PMA titers shown in **Figure 1** are visualized within the Spearman correlation matrix of HA0 and HA1 antigen-specific responses. Antibody binding data were $\log_{10}(x+1)$ transformed. Pairwise Spearman correlation coefficients were calculated and displayed as a correlation matrix, with purple indicating higher correlation coefficients and white indicating lower correlation coefficients. Abbreviations: PMA, protein microarray assay. Virus strain abbreviations can be found in **Supplemental Table 3**.

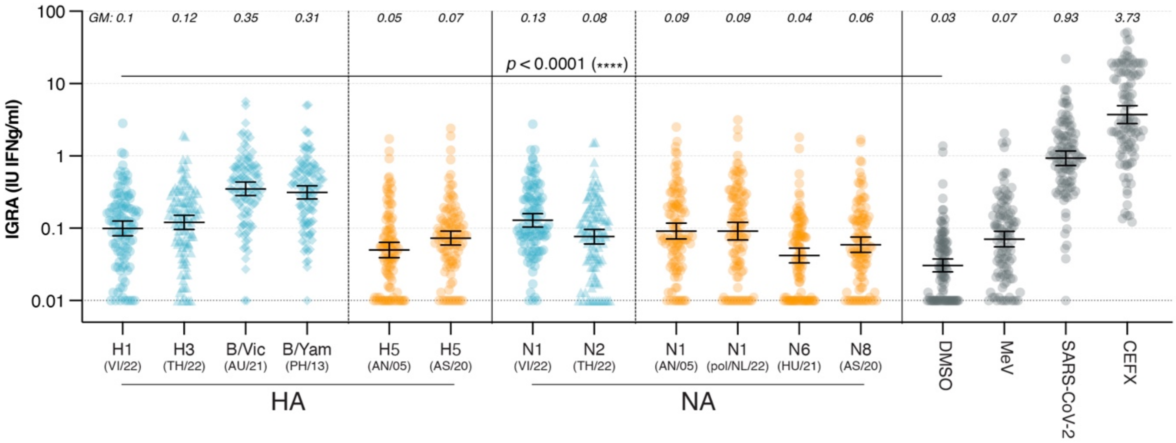


**Supplemental Figure 4. Uncorrected IFN-γ levels detected in IGRA.** IFN-γ levels (IU/mL) following stimulation of whole blood with overlapping HA or NA peptide pools representative of seasonal and avian influenza viruses. Measles virus N, SARS-CoV-2 S, and CEFX were included as controls. DMSO is shown as a background control. The bars indicate the GM with a 95% confidence interval. GM values for each peptide pool are listed above the scatter plot. Each dot represents an individual sample measurement from a total of 107 samples. Intra-individual responses to each HA and NA antigen were compared with the matched DMSO control using a paired, two-sided Wilcoxon signed-rank test. The resulting p-values were adjusted for multiple testing using the Benjamini–Hochberg procedure. All HA and NA antigen vs. DMSO comparisons were significant after correction (adjusted $p < 0.001$ for each HA and NA antigen). Abbreviations: IFN-γ, interferon gamma; IGRA, interferon-gamma release assay; IU, international units; MeV, measles virus; SARS-CoV-2, severe acute respiratory syndrome coronavirus-2; DMSO, dimethyl sulfoxide; GM, geometric mean. Virus strain abbreviations can be found in **Supplemental Table 3**.

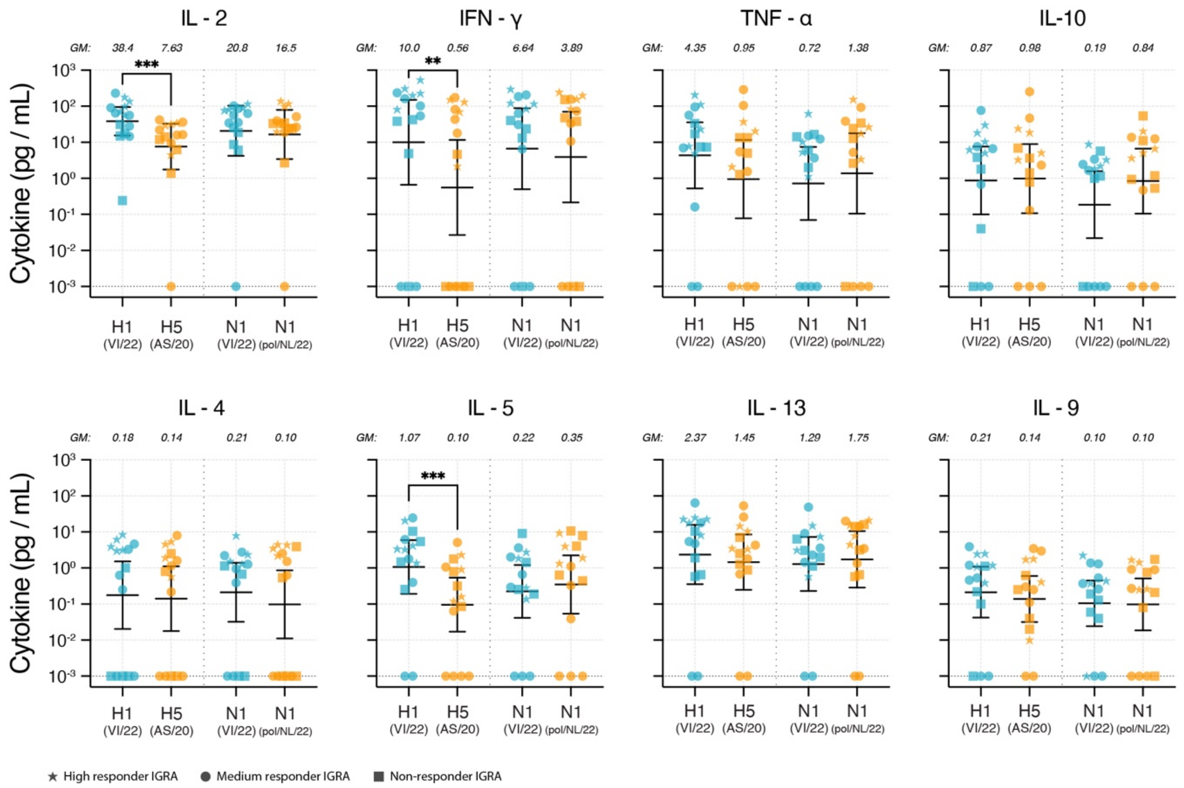


**Supplemental Figure 5. Detection of cytokine production in response to PBMC stimulation with overlapping HA and NA peptide pools in the supernatant of the AIM assay.** Cytokine levels were measured in AIM supernatants after stimulation with overlapping peptide pools covering HA and NA of seasonal (blue) or avian (yellow) influenza viruses. Levels of Th1 cytokines IL-2, IFN-γ, and TNF-α are displayed from left to right, followed by Th2 cytokines IL-10, IL-4, IL-5, and IL-13, and lastly, the Th9 cytokine IL-9. Each dot represents an individual sample measurement from a total of 15 individuals; DMSO background was subtracted. Bars indicate the GM with a 95% confidence interval. GM values for each peptide pool are listed at the top of each plot. The black horizontal line illustrates the detection limit for each cytokine shown. Differences between paired samples of either A(H1) VI/22 and A(H5) AS/20, or N1 VI/22 and N1 pol/NL/22 were evaluated using the Wilcoxon matched pairs signed rank test. NS, no significance; *, $p < 0.05$; **, $p < 0.01$; ***, $p < 0.001$; ****, and $p < 0.0001$. Abbreviations: PBMC, peripheral blood mononuclear cell; AIM, activation-induced marker; Th, T-helper; IL, Interleukin; IFN-γ, interferon gamma; TNF-α, tumor necrosis factor α; DMSO, dimethyl sulfoxide; GM, geometric mean. Virus strain abbreviations can be found in **Supplemental Table 3**.

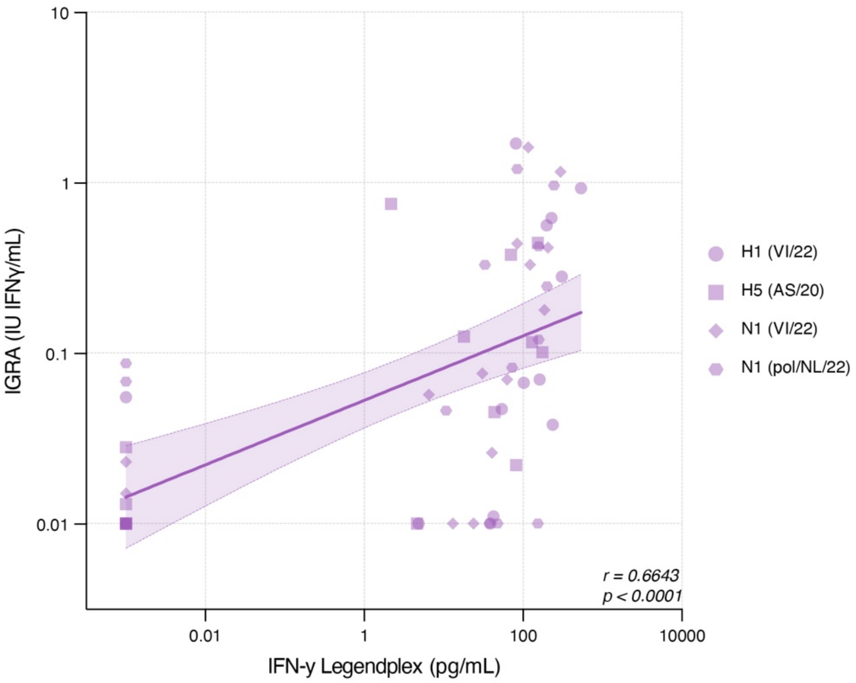


**Supplemental Figure 6. Correlation between IFN-γ responses measured by IGRA and AIM following influenza peptide stimulation.** IFN-γ levels were quantified after stimulation of PBMC with overlapping peptide pools representing seasonal A(H1) VI/22 and N1 VI/22, and avian A(H5) AS/20 and N1 pol/NL/22 influenza antigens. Data are shown for n = 15 individuals. IFN-γ levels from both assays were $\log_{10}$-transformed and plotted in correlation plots. Linear regression lines (solid) and 95% confidence intervals (shaded areas) are shown. Spearman correlation coefficients (r) and p-values are indicated. Abbreviations: IFN-γ, interferon gamma; IGRA, interferon-gamma release assay; AIM, activation-induced marker; PBMC, peripheral blood mononuclear cell.

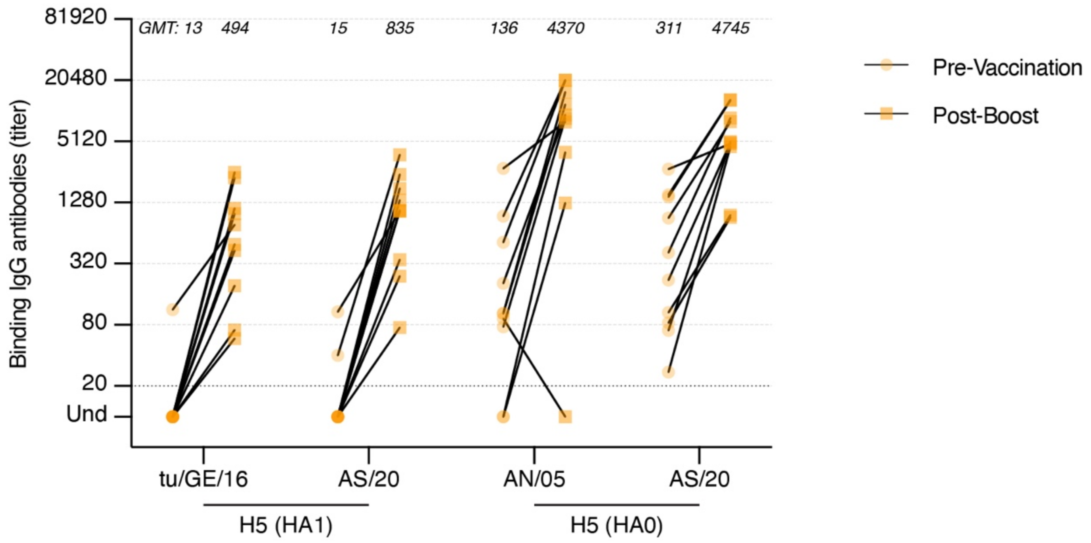


**Supplemental Figure 7. Detection of serum IgG binding antibodies by PMA to A(H5) influenza viruses pre- and post-A(H5) vaccination.** Serum samples were obtained in a previous A(H5) vaccination study conducted at the Erasmus Medical University Center and selected for validation purposes based on the presence of HI and neutralizing antibodies. Sera obtained pre-vaccination and post-booster vaccination were used. Binding IgG antibodies against a panel of HA1 and HA0 antigens from seasonal (A(H1), A(H2), A(H3), B(Vic), and B(Yam)) and avian (A(H5), A(H7)) influenza viruses were measured, binding IgG titers to relevant A(H5) antigens are shown in this figure. The dashed horizontal line represents the threshold titer of 20. Samples with unmeasurable PMA titers are indicated as undetectable. Abbreviations: PMA, protein microarray assay; HI, hemagglutinin inhibition; GMT, geometric mean titer. Virus strain abbreviations can be found in **Supplemental Table 3**.
